# CAR Density Influences Antitumoral Efficacy of BCMA CAR T cells and Correlates with Clinical Outcome

**DOI:** 10.1101/2022.01.19.22269515

**Authors:** Paula Rodriguez-Marquez, Maria E. Calleja-Cervantes, Guillermo Serrano, Aina Oliver-Caldes, Maria L. Palacios-Berraquero, Angel Martin-Mallo, Cristina Calviño, Marta Español-Rego, Candela Ceballos, Teresa Lozano, Patxi San Martin-Uriz, Amaia Vilas-Zornoza, Saray Rodriguez-Diaz, Rebeca Martinez-Turrillas, Patricia Jauregui, Diego Alignani, Maria C. Viguria, Margarita Redondo, Mariona Pascal, Manel Juan, Alvaro Urbano-Ispizua, Paula Rodriguez-Otero, Ana Alfonso-Pierola, Bruno Paiva, Juan Jose Lasarte, Susana Inoges, Ascension Lopez-Diaz de Cerio, Jesus San-Miguel, Carlos Fernandez de Larrea, Mikel Hernaez, Juan R. Rodriguez-Madoz, Felipe Prosper

## Abstract

Identification of new markers associated with long-term efficacy in patients treated with CAR T cells is a current medical need, particularly in diseases such as multiple myeloma. In this study we address the impact of CAR density on the functionality of BCMA-CAR T cells. Functional and transcriptional studies demonstrate that CAR T cells with high expression of the CAR construct show an increased tonic signaling with upregulation of exhaustion markers, increased *in vitro* cytotoxicity but a decrease in *in vivo* BM infiltration. Characterization of Gene Regulatory Networks using scRNA-seq identified regulons associated to activation and exhaustion upregulated in CAR^High^ T cells, providing mechanistic insights behind differential functionality of these cells. Finally, we demonstrate that patients treated with CAR T cell products enriched in CAR^High^ T cells show a significantly worse clinical response in several hematological malignancies. In summary, our work demonstrates that CAR density plays an important role in CAR T activity with significant impact on clinical response.

**Teaser:** High CAR molecule density affects CAR T cell activity and associates with impaired clinical response.

## INTRODUCTION

Chimeric antigen receptor (CAR) T cell therapies have emerged as a promising therapeutic tool against cancer, revolutionizing cancer immunotherapy (*1*). Second-generation CAR T cells have shown to induce impressive clinical responses in hematological malignancies, such as chemotherapy-resistant B cell leukemias and lymphomas (*2*–*4*) and multiple myeloma (MM) (*5*– *7*). Despite the high rates of remissions, not every patient achieves a complete response after CAR T-cell therapy. In addition, a significant number of patients experience a relapse of the disease. Particularly in patients with MM, despite their impressive responses, apparently, so far there is no a plateau in the survival curves after CAR T cell therapy (*5*–*7*), which contrast with results obtained with CD19 CAR T cells in acute lymphoblastic leukemia (ALL) and non-Hodgkin lymphomas (NHL).

It is well known that CAR T cells are heterogeneous products with multiple factors contributing to their efficacy. Several studies have demonstrated how extrinsic factors, like antigen density or tumor burden, strongly influence efficacy of CAR T cells (*8, 9*). In addition, factors related to CAR structure, like the costimulatory domain or the hinge length, can also affect the antitumoral potential of CAR T cells (*10*–*12*). Moreover, intrinsic cell factors, such as the differentiation state of T cells, the CD4/CD8 ratio or the T cell polyfunctionality have been correlated with the therapeutic efficacy and has led to the hypothesis that enrichment of CAR T cell products in T cells with a more immature phenotype may be associated with improvement in long term responses (*13*– *16*). On the other hand, an increase in T cells with an effector or exhausted phenotype may result in a reduced persistence of CAR T cells with a decrease in clinical response (*13*). Previous studies identified that CAR signaling in the absence of antigen stimulation, denominated tonic signaling, is associated to early T cell exhaustion (*17*–*20*), suggesting its role in triggering premature T-cell dysfunction. Additional factors related to the CAR construct may have an impact on functionality. For instance, recent studies have demonstrated that a more physiological expression of the CAR, by the integration of the transgene in the TRAC locus or the use of different promoters, can be associated with improved efficacy and reduced toxicity (*21*–*23*). These results also suggest that the density of CAR molecule in the membrane of CAR T cells might influence CAR signaling, affecting their antitumoral efficacy (*23, 24*). However, this hypothesis and the impact on clinical efficacy has not been formally explored.

Technological advances in genomics, such as single-cell sequencing, have allowed a notable progress towards understanding the genomic landscape of CAR T cells, providing some mechanistic insights into proper CAR T cell function (*25*–*29*). Moreover, functional commitment of CAR T cells is governed by complex Gene Regulatory Networks (GRN) that control CAR T cells at baseline as well as CAR T cell dynamics after antigen recognition being essential for CAR T functionality (*30*). Using scRNA-seq, recent studies have identified specific T cell signatures associated with efficacy and toxicity in patients with large B cell lymphomas (*26*), or molecular determinants of CAR T cell persistence such as *IRF7* mediated regulation of chronic interferon signaling (*29*), establishing this technology as a useful tool to improve efficacy of CAR T.

In this study, we address the impact of CAR density on the functionality of BCMA-CAR T cells. Phenotypic, functional, transcriptomic and epigenomic studies at bulk and single cell level revealed different profiles between CAR T cells with high and low expression of the CAR molecule (CAR^High^ and CAR^Low^ T cells). We show that CAR^High^ T cells are associated with tonic signaling and an exhausted phenotype, and identify the molecular mechanisms involved in the different functionality of CAR^High^ and CAR^Low^ T cells. We define a molecular signature associated with increase CAR density that, when applied to CAR T cell products, may predict clinical response. These results would provide a useful tool to understand the mechanisms behind proper CAR T cell function and identify biomarkers of response with potential clinical implications.

## RESULTS

### CAR T cells exhibit a wide range of CAR density on cell surface that influences CAR-mediated signaling

Current CAR T cell products are generated using retro/lentiviral vectors that render different levels of transduction and transgene expression within the cells, and consequently a wide range distribution in the number of CAR molecules in the surface of transduced cells. We hypothesized that this heterogeneity in CAR density could affect CAR-mediated signaling, and hence, influence the efficacy of CAR T cell products. To address this question, we first used a triple parameter reporter (TPR) system in Jurkat cells (*31*) to measure, by flow cytometry, CAR-mediated activation of the main signaling pathways (NFAT, NFkB and AP1) after tumor recognition using CAR T cells with different levels of CAR on the cell surface. Jurkat-TPR cells were infected with a second-generation CAR construct targeting BCMA, with 4-1BB as costimulatory domain, that was further modified to include a EGFRt reporter, facilitating the measurement of CAR level (Fig. S1). Then, subsets of Jurkat-TPR cells presenting different levels of CAR (termed CAR^High^ and CAR^Low^ cells) were selected according to the fluorescence intensity (FI) of EGFRt (top and bottom FI quartiles respectively, see methods), and analyzed after coculture with different BCMA-expressing MM cell lines (Fig. S1). We observed significantly increased levels of activation in the three mentioned pathways within the CAR^High^ population. Moreover, a significant increase of activation of CAR^High^ cells was also observed even in the absence of tumor cells, indicating an increase in tonic signaling at baseline (Fig. S1). We consistently observed this functional pattern with other CAR constructs targeting CD19, CD33 and HER2 (Fig. S1), indicating that a higher density of CAR molecules in the cell surface increases both the tonic signaling as well as the signal transduction after tumor encountering.

### CAR density influences antitumoral response of CAR T cells targeting BCMA

To further analyze the effect of CAR density on antitumoral efficacy, we characterized CAR T cells from ten healthy donors that were generated using a BCMA-targeting CAR construct (derived from ARI-0002h) co-expressing BFP as reporter marker (Fig. S2). CAR T cells were sorted into CAR^High^ and CAR^Low^ subpopulations based on the expression of BFP (Fig. 1A and S3). CAR^High^ T cells include those cells with a BFP FI > 1.2×10^4^ (average FI 26861 ± 5795), while the CAR^Low^ T cells subpopulation was restricted to BFP FI < 4×10^3^ (average FI 2513 ± 388). These FI values corresponded to the top and bottom FI quartiles respectively (Fig. S3). Then, BFP FI values were used to quantify the number of CAR molecules on the surface of these two CAR T cell subpopulations using an antibody-binding capacity bead assay. CAR^High^ T cells presented more than 5000 CAR molecules/cell, while the number of molecules/cell in CAR^Low^ T cells was below 1500 (Fig. S3). Vector copy number analysis revealed a significant higher number of viral integrations within CAR^High^ T cells, with an average of 4.5 ± 1.3 integrations in CAR^High^ T cells vs 1.8 ± 0.3 in CAR^Low^ T cells, and a significant increase in CAR mRNA expression levels was also observed, which could explain the increased CAR density observed in these cells (Fig. S3). We found increased cytotoxic activity and greater levels of IFN-γ, IL-2 and TNFα production in CAR^High^ T cells (Fig. 1B-C). To determine the translational value of these findings, we examined CAR T cell levels in CAR T cell products from an academic clinical trial (CARTBCMA-HCB-01; NCT04309981) (Fig. S3). We observed an increased *in vitro* lytic activity in those CAR T cell products enriched in CAR^High^ T cells (>30% of cells with >5000 CAR molecules/cell) (Fig. 1D).

**Fig. 1.**
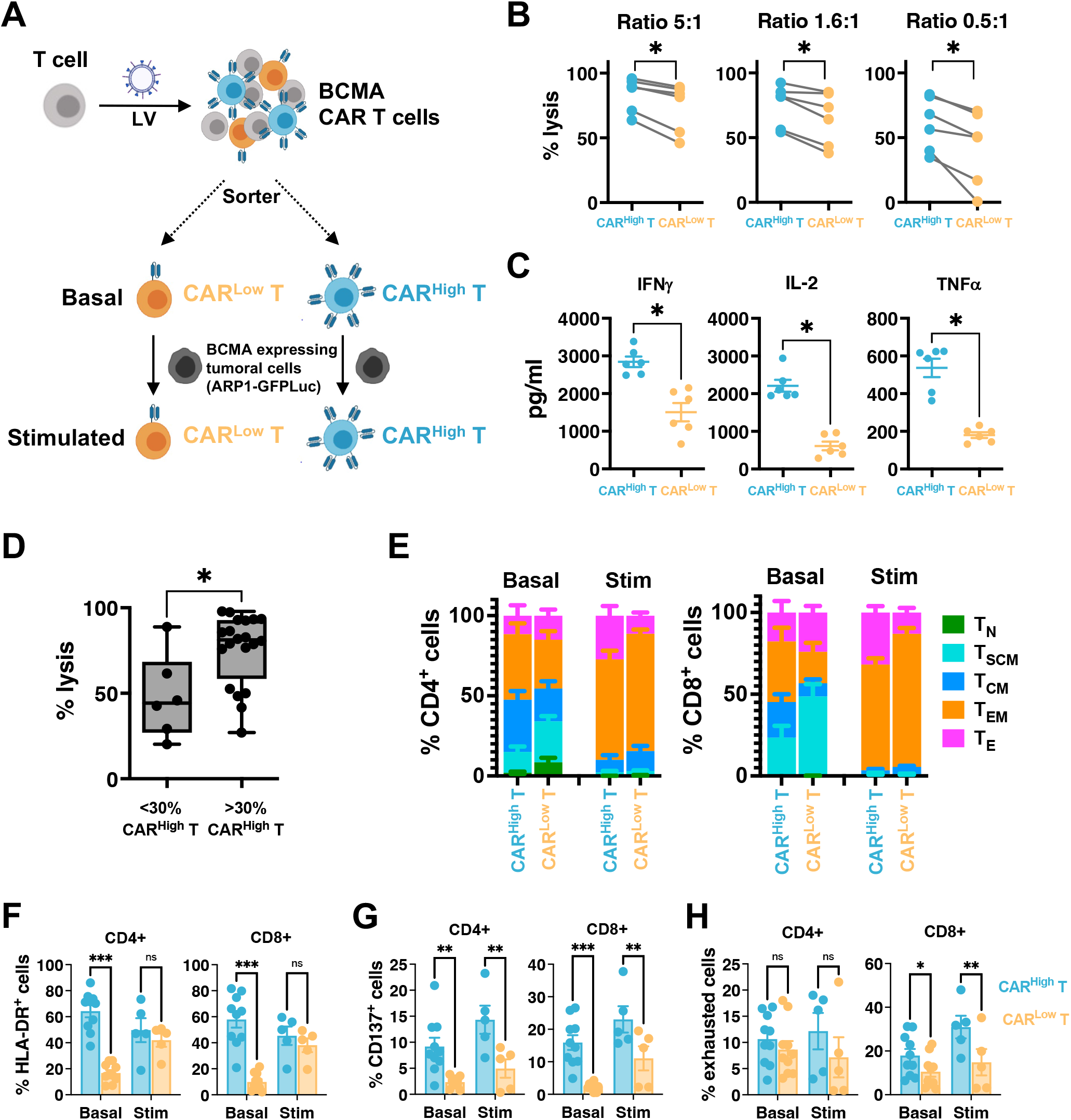
CAR^High^ T cells present increased *in vitro* antitumoral efficacy and exhausted phenotype. An *in vitro* functional and phenotypic characterization was performed on CAR T cells targeting BCMA presenting different densities of the CAR molecule. **(A)** Schematic representation of the procedure. CAR T cells were sorted into CAR^High^ and CAR^Low^ subpopulations via BFP expression. Analyses were performed at basal state or after stimulation with tumor cells expressing BCMA. **(B)** Quantification of the cytotoxic activity of CAR^High^ T and CAR^Low^ T cells against ARP1-GFPLuc at different E:T ratio. The percentage of specific lysis (average of three technical replicates) for each CAR T cell production (n=6) is depicted. **(C)** Quantification of IFNγ, IL-2 and TNFα levels in supernatants from cytotoxic assays (ratio 0.5:1) measured by ELISA. The cytokine concentration (pg/ml; average of three technical replicates) for each CAR T cell production (n=6) is depicted. **(D)** Cytotoxic activity of ARI-0002h CAR T products from CARTBCMA-HCB-01 clinical trial, according to their enrichment in CAR^High^ T cells. **(E)** Analysis of the phenotype of CAR^High^ and CAR^Low^ T cell populations before (basal n=10) and after stimulation (Stim n=5) with ARP1-GFPLuc tumor cells. Mean ± SEM of each T cell subpopulation within CAR^High^ T and CAR^Low^ T cells from is depicted. T_N_: naïve; T_SCM_: stem central memory; T_CM_: central memory; T_EM_: effector memory; T_E_: effector. Analysis of the expression of HLA-DR **(F)**, CD137 **(G)** and a combination of >2 exhaustion markers (LAG3, TIM3 and/or PD1) **(H)**, in CAR^High^ T and CAR^Low^ T cells before (basal n=10) and after stimulation (Stim n=5) with ARP1-GFPLuc tumor cells. Wilcoxon test for paired samples (B and C), Mann-Whitney test (D), and 2-way ANOVA with Sidak’s multiple comparison(F-H). *p<0.05; **p<0.01; ***p<0.001.

Next, we analyzed the phenotype of CAR^High^ and CAR^Low^ T cells before and after stimulation with tumor cells (Fig. 1E and S4). At baseline, we observed a statistically significant enrichment of central memory (T_CM_) and effector memory (T_EM_) phenotypes within CAR^High^ T cells, with concomitant reduction of naïve (T_N_) and stem central memory (T_SCM_) cells in both CD4^+^ and CD8^+^ subsets. After antigen stimulation, both populations acquired a T_EM_-T_E_ phenotype, although increased numbers of T_E_ were observed in CAR^High^ T cells (Fig. 1E and S4). These results suggest a higher degree of differentiation in CAR^High^ T cells even in the absence of antigen stimulation. Moreover, we observed that CAR^High^ T cells presented an increase in basal activation, with significant higher levels of HLA-DR^+^ and CD137^+^ cells, along with a higher percentage of CD8^+^ T cells expressing a combination of two or more markers of exhaustion (LAG3, TIM3 and/or PD1) (Fig. 1F-H). These differences in cell exhaustion increased after antigen stimulation, with more than 30% of CD8^+^ CAR^High^ T cells expressing an exhausted phenotype (Fig. 1H).

To determine whether different CAR densities would affect long-term antitumor potential, we evaluated their efficacy *in vivo* using a stress test in NSG mice (*21, 32*). ARP-1 cells expressing luciferase (1×10^6^ cells/animal) were transplanted in NSG mice and after 6 days 0.5×10^6^ CAR^High^ T or CAR^Low^ T cells FACS-sorted from a BCMA-CAR T cells were infused into the animals (Fig. 2A). Both subpopulations were able to eradicate tumor cells and increase survival of treated animals as demonstrated by luciferase measurements and by quantification of tumor cells into the bone marrow 28 days after treatment (Fig. 2B-E). However, when we analyzed the infiltration of CAR T cells in the bone marrow of the animals after 28 days, we observed a statistically significant reduction in the number of CAR T cells in animals treated with CAR^High^ T cells in comparison with CAR^Low^ T cells, suggesting a lower persistence of CAR^High^ T (Fig. 2F). Overall, these results indicate that CAR^High^ T cells show increased activation and tonic signaling and a more exhausted phenotype associated with an increase in *in vitro* cytotoxicity but a decrease in CAR T cell persistence in BM which could have an impact on the clinical response.

**Fig. 2.**
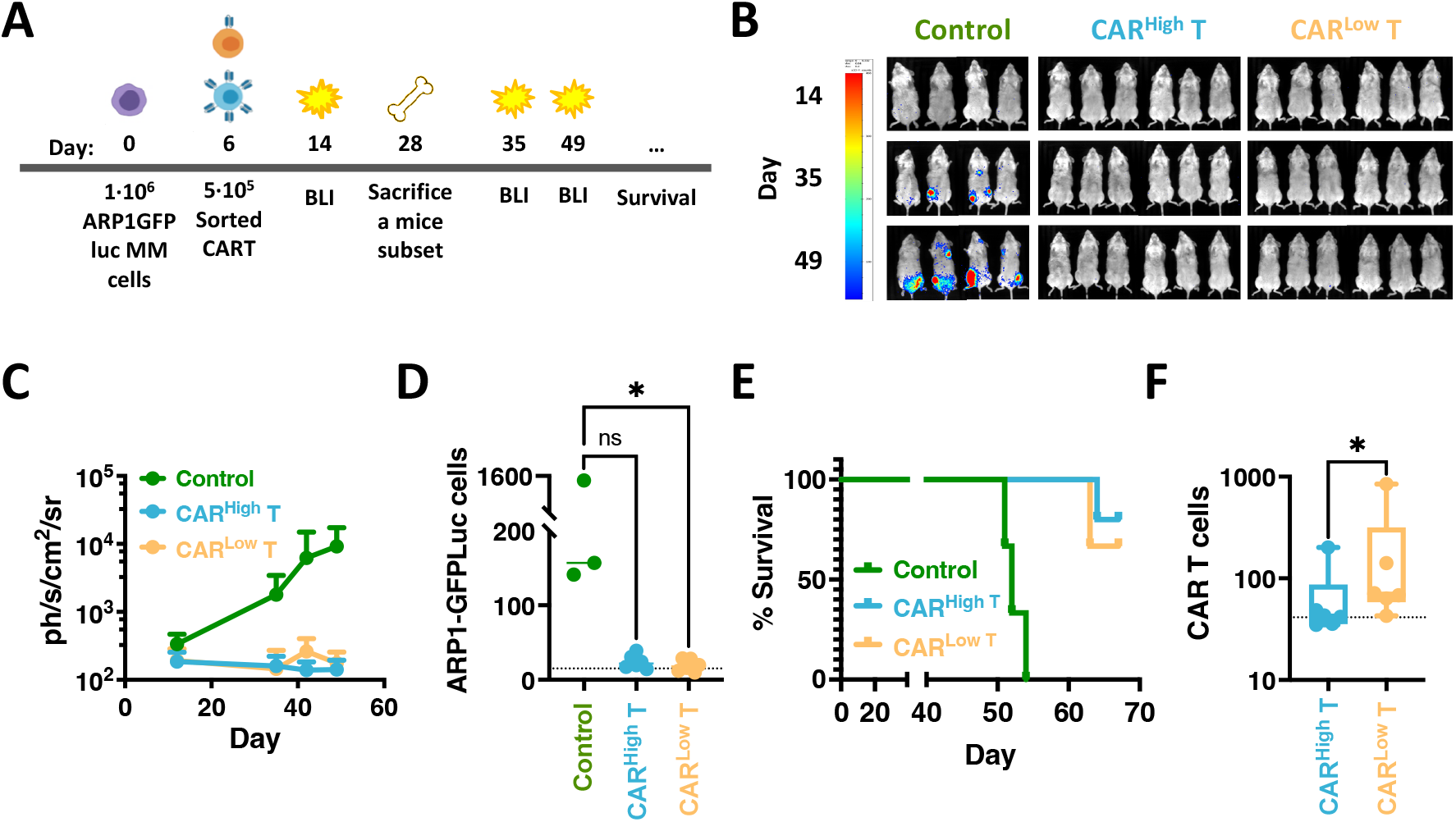
*In vivo* antitumoral efficacy of CAR T cells with different CAR densities. **(A)** Schematic representation of the experimental procedure. NGS mice were injected intravenously (i.v.) on day 0 with 1×10^6^ ARP1GFPLuc cells/animal. After 6 days 0.5×10^6^ CAR^High^ T or CAR^Low^ T cells were injected i.v. Bioluminiscence analysis (BLI) was performed on days 14, 35 and 49. Animal survival was monitored until the end of the experiment (day 67). On day 28 a subset of animals was sacrificed to analyze the presence of tumor and CAR T cells. **(B)** BLI images at the indicated days of control mice (n=4) or treated with CAR^High^ T cells (n6) or CAR^Low^ T cells (n=6). **(C)** Quantification of BLI (ph/s/cm^2^/sr) as a measurement of tumor growth. **(D)** Quantification of the tumor cells present in the bone marrow (BM) of control mice (n=3) or treated with CAR^High^ T cells (n=6) or CAR^Low^ T cells (n=6) at day 28 after CAR T cell administration. **(E)** Survival of control mice (n=4) or treated with CAR^High^ T cells (n=6) or CAR^Low^ T cells (n=6). **(F)** Quantification of the CAR T cells present in the BM of animals treated with CAR^High^ T cells (n=6) or CAR^Low^ T cells (n=6) at day 28 after CAR T cell administration. Kruskall-Wallis test (D), Mantel-Cox (Long-Rank) test (E), and Mann-Whitney test (E). ns: not significant; *p<0.05.

### CAR^High^ T cells display different transcriptomic and chromatin landscape with increased tonic signaling and T cell activation

Given the differences observed in phenotype and persistence between CAR^High^ T and CAR^Low^ T cells, we delved into the transcriptomic and epigenetic landscape of the two subpopulations of CAR T cells. Sorted CD4^+^ and CD8^+^ CAR^High^ and CAR^Low^ T cell populations from six different CAR T cell productions were profiled using high-throughput RNA sequencing (RNA-seq) and assay for transposase-accessible chromatin (ATAC-seq). Transcriptomic analysis revealed less than 100 genes differentially expressed (DEGs, FDR<0.05, Log2FC >2) between CAR^High^ T and CAR^Low^ T (Table S1). Similarly, the analysis of the ATAC-seq data revealed: i) a similar peak distribution in both populations, with >60% of the peaks located within the promoter regions and the first intron, and ii) a limited number of differential accessible regions identified between CAR^High^ T and CAR^Low^ T (Fig. S5 and Table S2). Interestingly, these small transcriptomic and epigenomic differences were enough to separate CAR^High^ T cells from CAR^Low^ T cells in a principal components analysis in both CD4^+^ and CD8^+^ T cell subsets (Fig. 3A and S6). Interestingly, DEG between CAR^High^ T and CAR^Low^ T cells were associated to genes involved in tonic signaling and T cell activation (Fig. 3B and S6). These results corroborated our phenotypic observations (see previous sections), where CAR^High^ T cells showed increased expression level of genes related to lymphocyte activation, such as HLA-DR or CD74, and costimulatory molecules, as TNFRSF4 (OX40) and TNFRSF9 (4-1BB) (Fig. 3C and S6). Moreover, these results were consistent with the results observed in Jurkat-TPR experiments (see section above). While reduced overlapping was observed between DEGs and differential peaks, those genes showing differential expression and chromatin accessibility were mainly related to the activation and tonic signaling, including HLA-DRA, CIITA, TNFRSF9 and CTLA-4 (Fig. 3D and S6). Together, these results suggest that small differences in gene expression and chromatin accessibility associated to increased CAR levels can substantially modify the overall phenotypic and functional profile of CAR T cells.

**Fig. 3.**
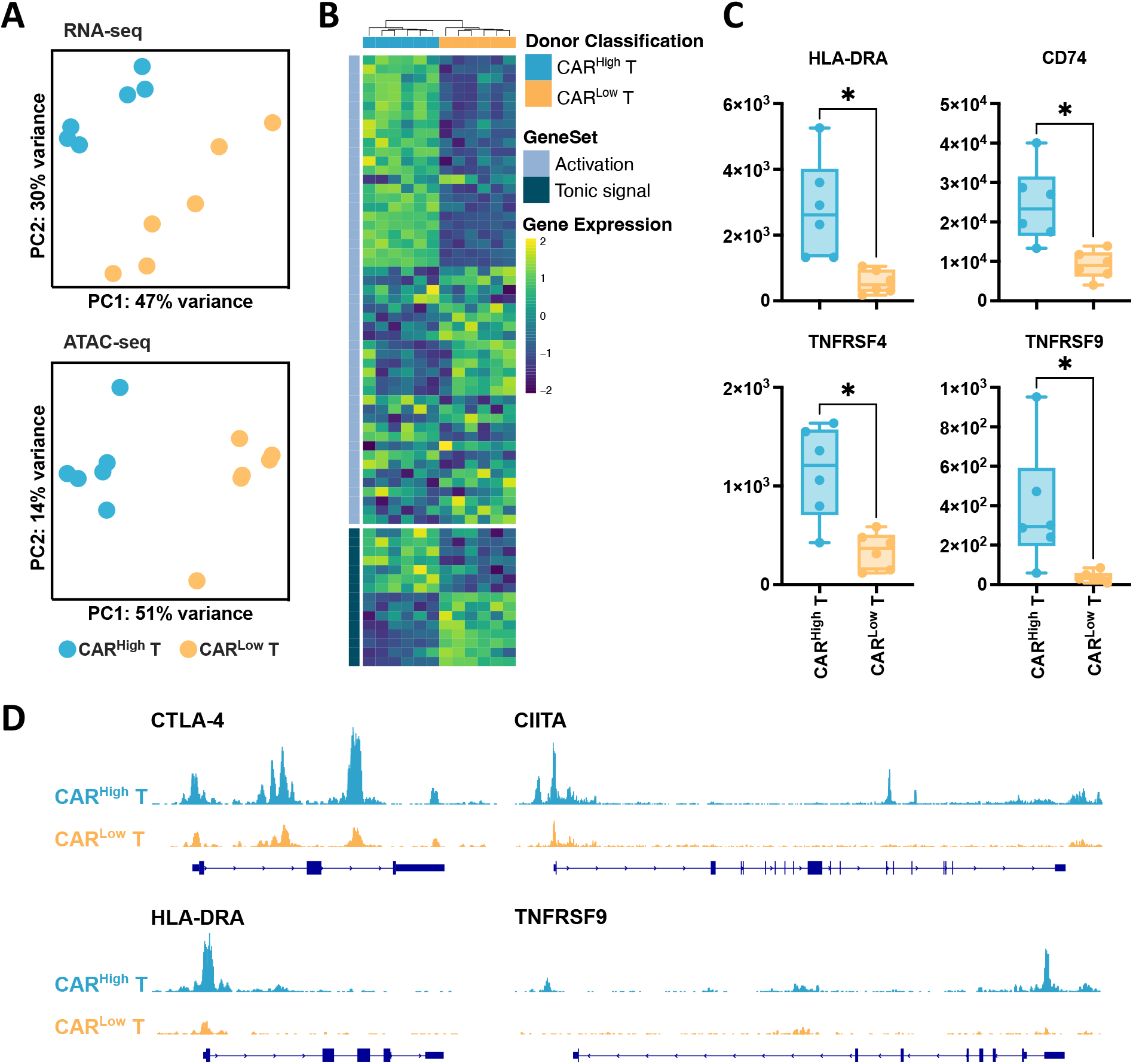
Transcriptomic profile and chromatin landscape of CD8^+^ CAR^High^ T cells. The transcriptomic and epigenetic landscape of sorted CD8^+^ and CD4^+^ (see Fig. S6) CAR^High^ and CAR^Low^ T cells (n=6) were profiled using high-throughput RNA sequencing (RNA-seq) and assay for transposase-accessible chromatin (ATAC-seq). **(A)** RNA-seq and ATAC-seq principal components (PC) analysis, corrected by patient heterogeneity, of sorted CD8^+^ CAR T cell subsets. Percentage of variance explained by PC1 and PC2 are depicted. **(B)** Heatmap of differentially expressed genes (DEGs) between CD8^+^ CAR^High^ T and CAR^Low^ T cells associated to genes involved in tonic signaling and T cell activation. **(C)** Quantification of HLA-DR, CD74, TNFRSF4 (OX40) and TNFRSF9 (4-1BB) gene expression in CD8^+^ CAR^High^ T and CAR^Low^ T cells. **(D)** UCSC genome browser tracks of CTLA-4, HLA-DRA, CIITA and TNFRSF9 showing differential peaks from ATAC-seq analysis between CD8^+^ CAR^High^ T and CAR^Low^ T cells. Wilcoxon test for paired samples (C). *p<0.05.

### Single-cell sequencing reveals specific distribution of CAR^High^ T cells

To better understand the heterogeneity of CAR T cells and the influence of CAR level on their transcriptomic profile, we performed single cell transcriptomic analysis on 43,981 CAR T cells from three independent productions. After quality control and filtering, we performed an integrated analysis and identified 23 clusters of CAR T cell subpopulations (Fig. S7). Clusters identified based on cell cycle gene signatures, those containing high levels of mitochondrial genes, as well as clusters lacking expression of T cell markers were excluded from further analysis (Fig. S7). Cell types and functional states of the remaining 15 clusters (containing 28,117 cells with range of 7,416-10,5093 cells/donor) were defined according to the expression of previously described canonical markers (*26*–*28*) (Fig. 4A-B). Within CD4^+^ cells we identified early memory (IL7R), memory (TCF7, CCR7, CD27), activated (HLA-DRA, OX40), cytotoxic (GZMA, PRF1) and Th2 helper (GATA3) CAR T cells, with a minority of other subtypes including cells expressing genes related to glycolysis and IFN response. Among CD8^+^ cells, we distinguished memory (TCF7, CCR7) and cytotoxic (GZMA, PRF1, NKG7) CAR T cells. Furthermore, in accordance with the phenotypical results, we identified a pre-exhausted cluster of CD8^+^ CAR T cells characterized by the expression of cytotoxic genes along with inhibitory receptors, like LAG3 and TIGIT (*33*) (Fig. 4B-C). Analysis of V(D)J rearrangement showed a polyclonal diversity of CAR T cells within all the clusters, indicating no specific enrichment of any particular clone (Table S3).

**Fig. 4.**
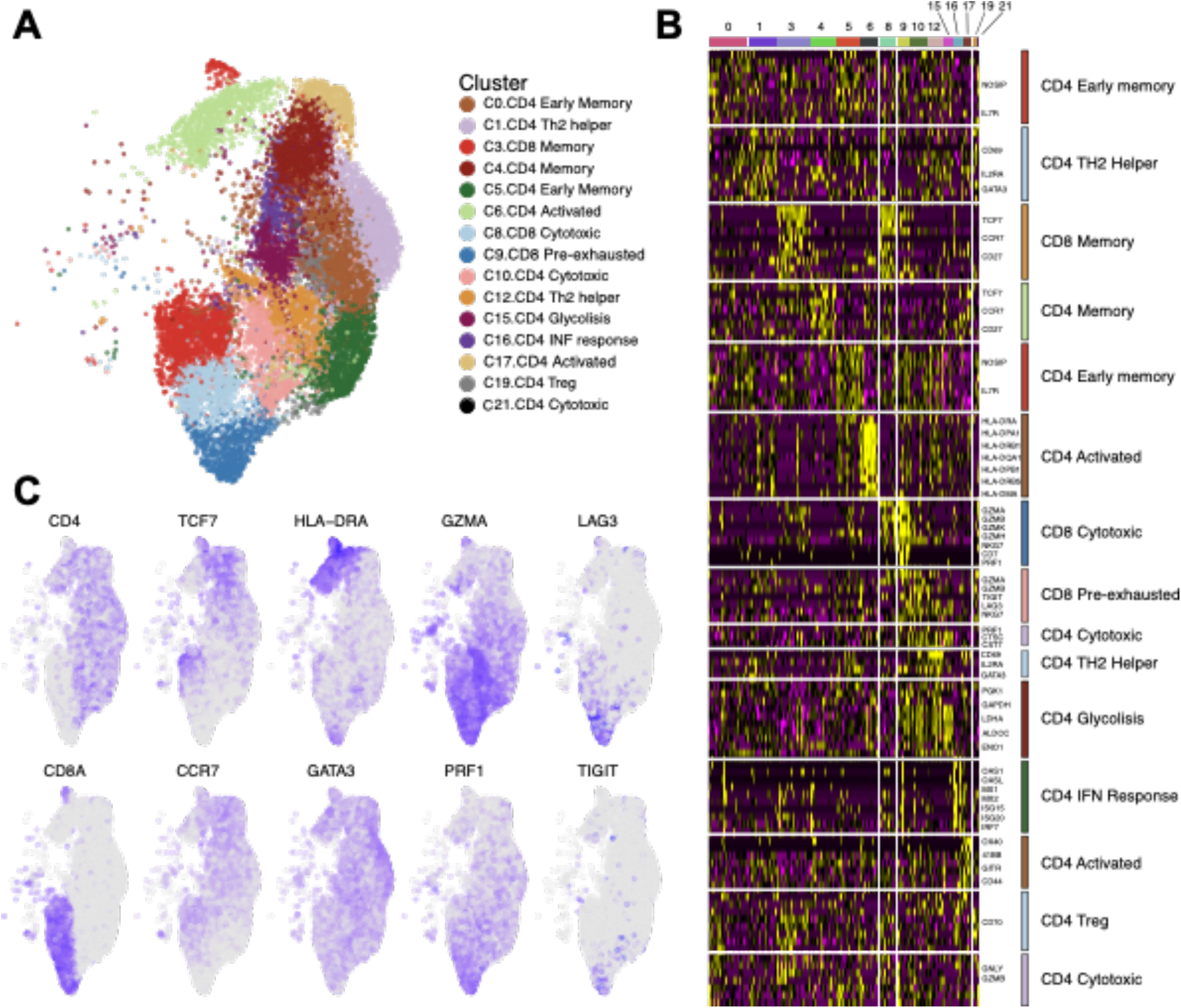
Characterization of CAR T cells at single cell level. CAR^+^ (BFP^+^) cells from three independent CAR T cell productions were assayed by single-cell RNA sequencing. **(A)** An overview of the 28,117 cells that passed QC and filtering for subsequent analyses in this study. UMAP plot showing the 15 clusters that were analyzed. **(B)** Heatmap showing signature genes of each cluster and putative assignments to cell types according to canonical marker genes. **(C)** UMAP plot overlaid with mRNA expression of T cell markers (CD4 and CD8A), memory markers (TCF7 and CCR7), activation markers (HLA-DRA and GATA3), effector enzymes (GZMA and PRF1) and exhaustion markers (LAG3 and TIGIT).

To robustly identify CAR T cells with high expression of the CAR construct in our single cell data, we developed gene signatures associated to both CD4^+^ and CD8^+^ CAR^High^ T cells, using DEGs from bulk RNA-seq analysis between CAR^High^ T and CAR^Low^ T cells (Table S4), as these populations were sorted based on CAR protein expression (see methods). We found a better correlation between the gene signature and CAR protein level, than the one observed between the expression of the CAR gene and its protein expression (Fig. S8). Moreover, the possibility to extrapolate these gene signatures to other datasets was assessed via leave-one-out cross-validation (Fig. S8). Then, we annotated CAR^High^ T cells in our single cell data using the developed signatures (Fig. 5A), and we observed that CAR^High^ T cells mainly localized within activated CD4^+^ cells, representing more than 50% of the cells in cluster 6 and almost 80% of the cells in cluster 17 (Fig. 5B). Furthermore, in the CD8^+^ T cell compartment, CAR^High^ T cells were more represented in cluster 9 showing a pre-exhausted phenotype (Fig. 5B). These results are in accordance with our previous phenotypic and transcriptomic analysis performed in sorted CAR^High^ T and CAR^Low^ T cell populations, where CAR^High^ T cells were significantly enriched in activation and tonic signaling signatures (Fig. 5C-D). Moreover, we perform this analysis using an independent single cell public dataset of CAR T products targeting CD19 from patients with DLBCL (*26*), obtaining similar results (Fig. S9). Altogether, our results would indicate a strong association between tonic signaling, CAR T cell activation and cell exhaustion that is increased in CAR T cells with high CAR density defined either by the gene signature or the protein expression.

**Fig. 5.**
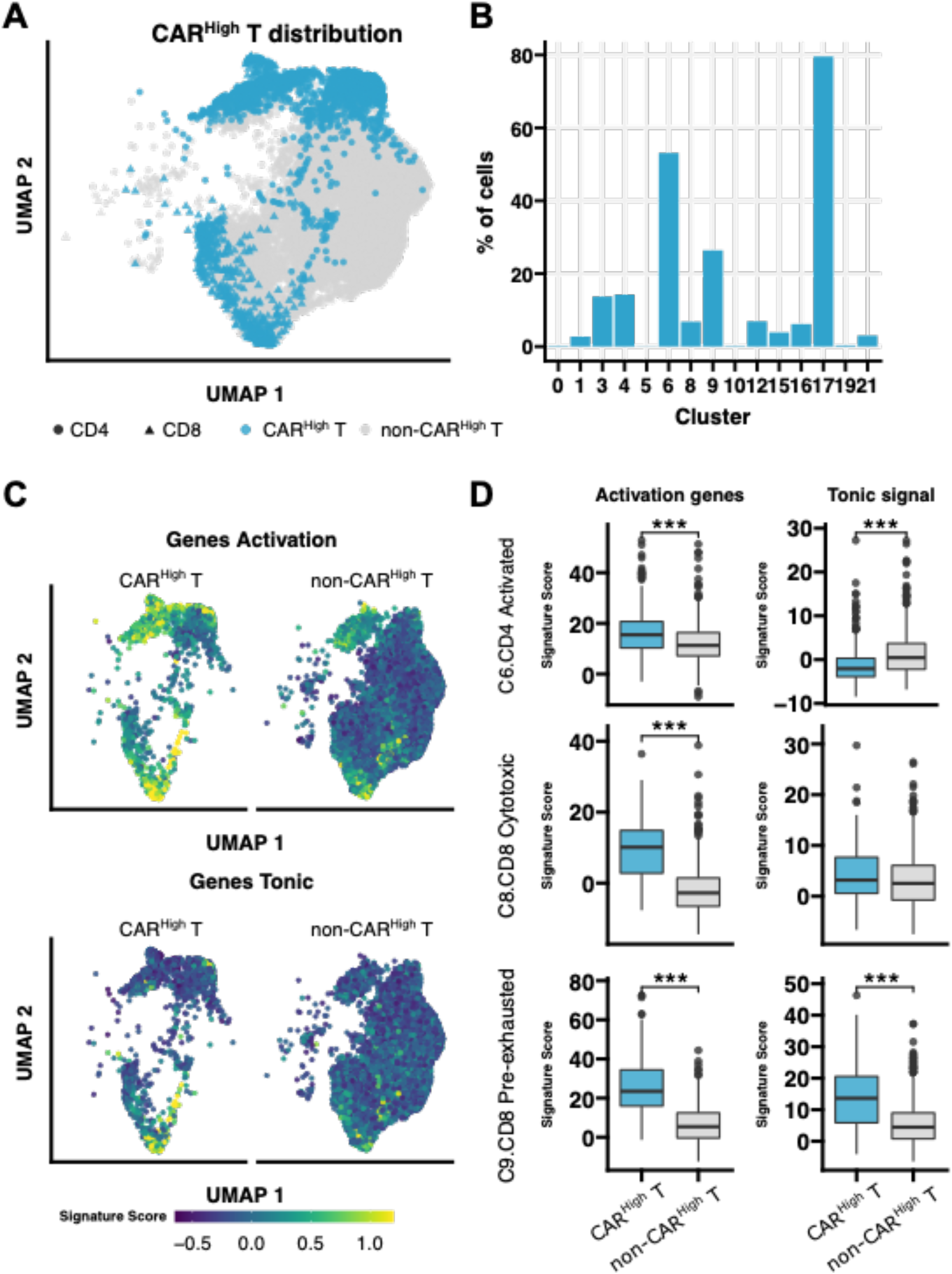
Single-cell sequencing reveals specific distribution of CAR^High^ T cells. Annotation and further analysis of CAR^High^ T in the single cell data was performed by applying the gene signatures, developed in this work, associated to both CD4^+^ and CD8^+^ CAR^High^ T cells, that showed higher correlation with the CAR protein level than that yielded by the CAR gene expression. **(A)** UMAP plot showing CAR^High^ T cell distribution across analyzed cells. **(B)** Quantification of the percentage of CAR^High^ T cell along the different clusters. In accordance with phenotypic results, CAR^High^ T cells are mainly localized within activated CD4^+^ cells (cluster 6 and 17) and pre-exhausted CD8^+^ T cells (cluster 9). **(C)** UMAP plots overlaid with the score of activation and tonic signaling signatures, showing their distribution across cells. CAR^High^ T cells were enriched in the score for both signatures. **(D)** Quantification of the signature score in CAR^High^ T cells from clusters 6 (CD4^+^ activated), 8 (CD8^+^ cytotoxic) and 9 (CD8^+^ pre-exhausted), in comparison with the rest of the cells, for both activation and tonic signaling signatures. CAR^High^ T cells presented a significant increase for both signatures in almost all three clusters analyzed. Wilcoxon test (D). ***p<0.001.

### CAR density is associated with differential activation of regulatory networks

To elucidate the molecular regulation of CAR^High^ T cells we applied SimiC (*34*), a novel GRN inference algorithm for scRNA-seq data that imposes a similarity constraint when jointly inferring the GRNs for each specific cell state. Based on this analysis, we observed regulons (a TF and its associated target genes) that were similarly activated between CAR^High^ and the rest of the CAR T cells (Fig. S10), such as regulons implicated in T cell differentiation (GATA3, RUNX3) and signal transduction (STAT1, REL, RELA, JUN/AP1) (*35*–*37*). On the other hand, we identified some regulons that were more active in CAR^High^ T cells, like STAT3, a TF associated with development and maintenance of T cell memory (*38*), or ARID5A, a TF related to the control of the stability of STAT3 (*39*) (Fig. S10) and other regulons that presented reduced activity in CAR^High^ T cells, like BTG2, a TF related to the prevention of proliferation exacerbation and spontaneous activation (*40*) (Fig. S10). These changes in regulon activity could explain the increased central memory phenotype and also provide a regulatory mechanism for the increased activation observed in CAR^High^ T cells.

Interestingly, we also observed regulons presenting a multimodal activation profile. To determine whether this distribution might be related to different activity between clusters, we computed the distribution of the regulon activity in each cluster (provided it contain at least 5% of CAR^High^ T cells) (Fig. S10). As an example, we observed that the activity of RFX5 regulon, a member of the RFX family that interacts with HLA class II genes and promotes their transcription (*41, 42*), was overexpressed in CD8^+^ CAR^High^ T cells, independently of the T cell subtype. However, RFX5 regulon activity progressively increased through CD8^+^ differentiation when we analyzed CD8^+^ CAR T cells that were not CAR^High^ T, from memory, to cytotoxic and finally to pre-exhausted cells (Fig. 6A). In addition, we searched for regulons that could explain the exhausted phenotype observed in CAR^High^ T cells. We observed that NR4A1 and MAF regulons, already described as drivers of T cell exhaustion (*43*–*45*), were more active in CAR^High^ T cells (Fig. 6A). On the other hand, the SATB1 regulon, related to PD1 inhibition (*46*), presented lower activity in CAR^High^ T cells (Fig. 6A). All these results may provide molecular mechanism underpinning the exhausted phenotype observed in CAR^High^ T. Overall, our GRN analysis using SimiC provides mechanistic insights into the regulatory networks behind the phenotypic and functional differences observed in CAR^High^ T cells, identifying regulons that regulate T cell function previously described, supporting the usefulness of this methodology. More importantly, the use of SimiC permits the generation of novel hypothesis based on identified regulatory factors that could be modulated, ultimately to the design of optimized CAR T therapies.

**Fig. 6.**
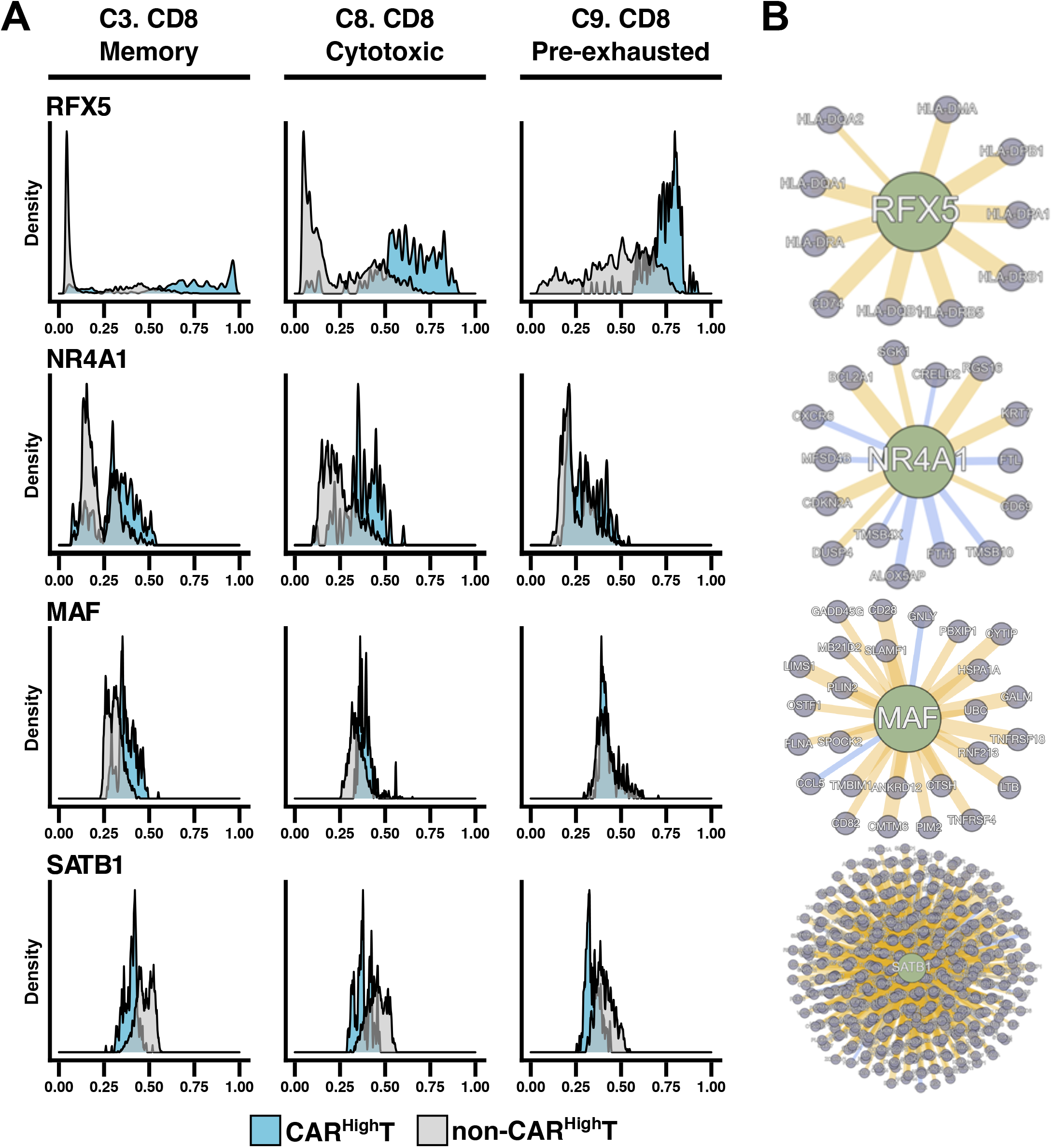
CAR density is associated with differential activation of regulatory networks. Dynamics in Gene Regulatory Networks (GRN) of CAR^High^ T cells were analyzed by SimiC, a novel GRN inference algorithm for scRNA-seq data that imposes a similarity constraint when jointly inferring the GRNs for each specific cell state. Histograms showing the activity score during CD8^+^ T cell differentiation in CAR^High^ T cells versus the rest of the cells **(A)**, and the inferred gene network **(B)**, of regulons RFX5, NR4A1, MAF and SATB1. Regulon activity of RFX5, a member of the RFX family that interacts with HLA class II genes and promotes their transcription, and NR4A1 and MAF, that have been described as drivers of T cell exhaustion, was already high in CAR^High^ T cells, independently of the phenotype, meanwhile in the rest of the cells, their activity progressively increased, from CD8^+^ memory (cluster 3), to CD8^+^ cytotoxic (cluster 8) and finally to CD8^+^ pre-exhausted (cluster 9) phenotypes. In contrast activity of SATB1 regulon, related to PD1 inhibition, was reduced in CAR^High^ T cells.

### CAR^High^ T gene signature is associated with clinical response

Given the differences in functionality between CAR^High^ and CAR^Low^ T cells, we reasoned that CAR density might have an impact on the clinical response to CAR T-cell therapies. To evaluate this hypothesis, we applied the gene signatures associated with CAR^High^ T cells to infusion products from several clinical trials with public transcriptomic data available (*25, 26*). We first applied our CD4^+^ and CD8^+^ signatures to bulk RNA-seq data of 34 infusion products from adult CLL patients treated with CTL019 (*25*). We observed that products from patients with poor clinical response (partial responders and non-responders) presented a significant higher score of both CD4^+^ and CD8^+^ CAR^High^ T signatures (Fig. 7A). We also assessed CAR^High^ T signature on a single cell RNA-seq dataset comprising anti-CD19 CAR T infusion products from 24 patients with DLBCL (*26*). We found that the products from non-responder patients were significantly enriched in CD8^+^ CAR^High^ T cells (Fig. 7B), supporting the correlation between CAR density and clinical response. Finally, we examined the correlation between the clinical response and the expression of CAR measured by FACS, in the cell products of an academic clinical trial assessing ARI-0002h, a CAR T cell targeting BCMA (CARTBCMA-HCB-01; NCT04309981). Patients with partial response (less or equal than VGPR) showed an increase in the number of CAR^High^ T within the infusion product versus the patients presenting sCR (Fig. 7C). Moreover, a shorter duration of response was observed in patients with increased percentage of CAR^High^ T cells (p=0.04, as continuous variable in Cox regression model). No statistical correlation was observed with the development or grade of cytokine release syndrome (CRS), although the patients of this cohort presented only low-grade CRS (grades 1 and 2). Overall, our data suggest that CAR T therapeutic products enriched in T cells with high CAR density on the membrane, would impact negatively on the clinical response.

**Fig. 7.**
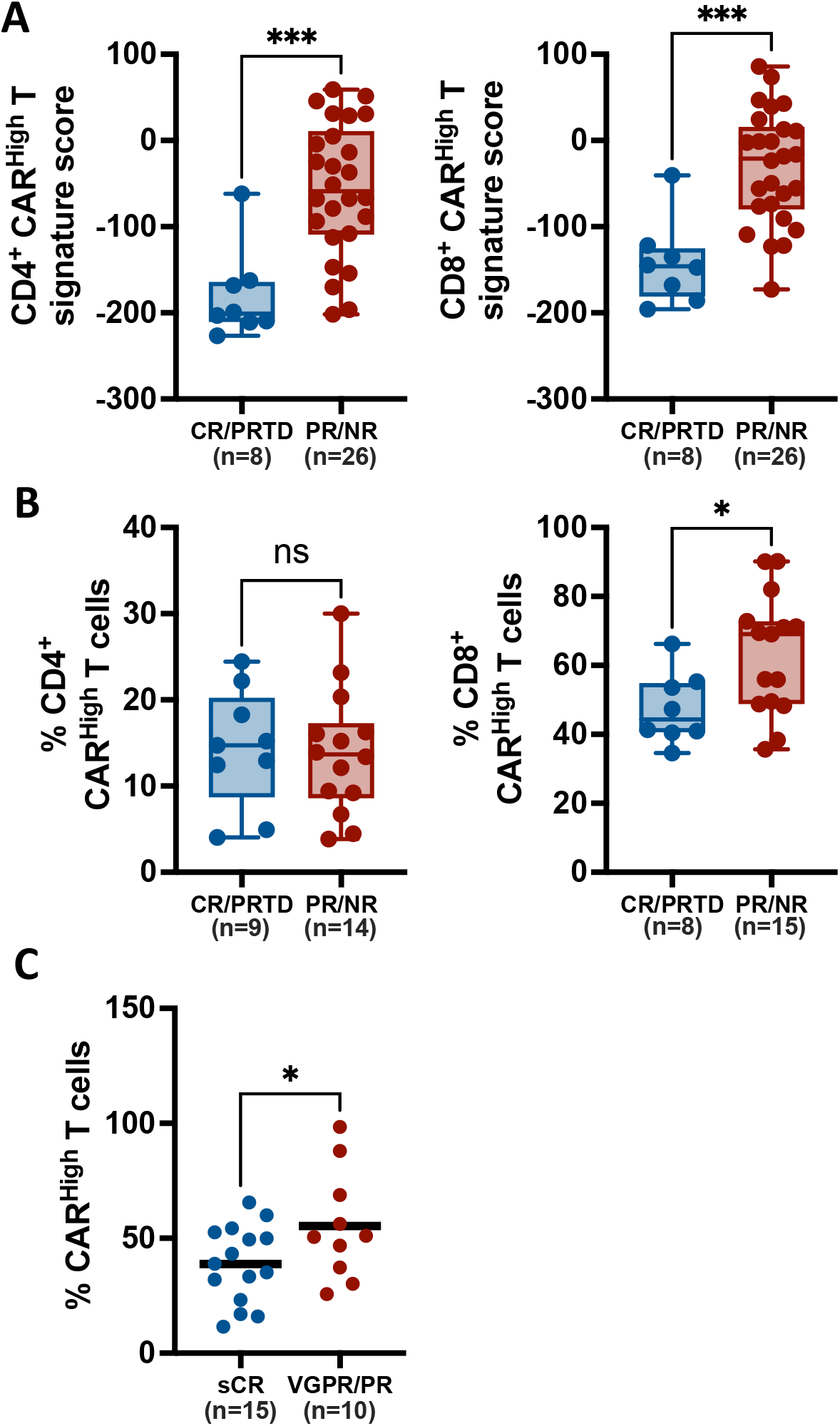
Increased CAR levels negatively impact clinical response of CAR T therapies. To evaluate the impact of increased CAR density on the clinical response, we quantify the presence of CAR^High^ T cells in the infusion products of several clinical trials. **(A)** The gene signatures developed in this works, associated to CD4^+^ and CD8^+^ CAR^High^ T cells, were applied to 34 infusion products from adult CLL patients treated with CTL019. Signature score for each CAR T cell product is represented for both CD4^+^ (left) and CD8^+^ (right) gene signatures, in patients divided according to clinical response into CR/PRTD (Complete Response/Partial Response with Transformed Disease) and PR/NP (Partial Response/Non Response). Products from patients with poor clinical response (PR/NR) presented a significant higher score of both CAR^High^ T signatures. **(B)** Our gene signature was applied to available single cell RNA-seq dataset from 24 CAR T infusion products from adult DLBCL patients treated with axi-cel. Percentage of CD8^+^ CAR^High^ T present for each CAR T cell product is represented in patients divided according to clinical response as described in (A). Products from PR/NR patients were significantly enriched in CD8^+^ CAR^High^ T cells. **(C)** The number of CAR molecules/cell in 25 anti-BCMA CAR T infusion products from CARTBCMA-HCB-01 clinical trial was quantified by FACS. CAR^High^ T cells were defined as these cells with >5000 molecules/cell (see methods). Percentage of CAR^High^ T cells for each infusion product is represented in patients divided into sCR (Stringent Complete Response) and VGPR/PR (Very Good Partial Response/Partial Response) according to clinical response. Patients with sCR showed a significantly decreased number of CAR^High^ T cells. Unpaired t tests (A-C). *p<0.05; ***p<0.001.

## DISCUSSION

The functionality of CAR T cells relies on the interaction between the tumor and engineered T cells (*1*). However, as living drugs, CAR T cells are heterogeneous products in which intrinsic and extrinsic factors can influence their functionality having a significant impact on their clinical efficacy (*8*–*16*). Among others, the level of antigen expression has been associated with anti-tumor response (*8, 24*). Our study contributes to identify new determinants of CAR T cell function demonstrating a clear role of the level of CAR expression on the functionality of CAR T cells. Our results indicate that high levels of CAR expression are associated with increased tonic signaling and a cell exhausted phenotype, characterized by the expression of multiple inhibitory receptors, such as PD1, CTLA4, LAG3, TIM3 and TIGIT, among others. This phenotype has been associated to reduced responses and worse long-term relapse free survival (*13, 25, 48*), which is consistent with our findings demonstrating decreased responses in patients with increased levels of CAR^High^ T cells in different hematological malignancies.

Previous studies have demonstrated that constitutive signaling induced by multiple factors related to the different CAR moieties (*49, 50*), are associated to early T cell exhaustion (*17*–*20*). For instance, a recent study has shown fundamental differences in CAR signaling between CAR T cells with CD28 or CD8 transmembrane domains (TMD) related to the heterodimerization potential of the different transmembrane domains (*51*). Our results indicated that an increased density of the CAR molecule in the surface of the T cells (CAR^High^ T cells), triggered by a higher number of viral integrations, could be enough to induce spontaneous clustering of the chimeric receptor molecules leading to tonic signaling. Interestingly, tonic signaling was not restricted to CAR^High^ T cells targeting BCMA, since it was also observed in CAR^High^ T cells with different specificities (CD19, CD33 and HER2), suggesting that this phenomenon was related to CAR density rather than CAR specificity.

Most CAR T constructs are generated using retro/lentiviral vectors with strong promoters, like human EF1a or the murine stem cell virus LTR (MSCV), with no physiological regulation. These promoters induce high levels of CAR expression that can lead to tonic signaling and premature exhaustion. The possibility of reducing gene expression via weaker promoters as well as through more physiological promoters has been associated with decreased tonic signaling (*22, 23, 52*). In fact, the use of the MND promoter has been shown to reduce CAR surface density, while EF1a promoter increased its density, leading to a higher cytotoxic activity, cytokine production and expression of exhaustion markers (*23*). Thus, the use of physiological promoters would be a strategy to prevent an increase in CAR^High^ T cells, thus limiting exhaustion of CAR-T and favoring long term persistence and antitumor efficacy (*22*).

The increased tonic signal and the concomitant basal activation of CAR^High^ T cells resulted in an increase in tumor cytotoxicity *in vitro*. The higher cytotoxicity and cytokine production observed in CAR^High^ T cells is also consistent with previous reports describing differences in antitumoral efficacy due to increased ligand-independent signaling (*32, 53*). Nevertheless, the pre-exhausted phenotype of CAR^High^ T cells could compromise their long-term persistence and function. Our results from the *in vivo* experiments based on “stress tests” (*32, 54*) support this hypothesis, since we observed reduced bone marrow infiltration of CAR^High^ T cells. However, the use of CAR^Low^ T cells was not associated with improved survival of the animals since both CAR T populations were able to induce strong antitumor responses. Unfortunately, the use of immunodeficient mouse models is a significant limitation for testing CAR T cell efficacy, as well as toxicity. The use of humanized mice, or syngeneic mouse models together with mouse CAR T cells, should help to better characterize *in vivo* the impact of CAR expression of long-term efficacy of the different products. Nevertheless, the clinical correlation between response and decrease number of CAR^High^ T cells in the infusion product would also be consistent with the hypothesis that infusion of CAR^High^ T cells could compromise their long-term persistence and function.

The avidity of the scFv against the antigen plays an important role on the functionality of the CAR T cells and its control may result in more effective therapies (*55, 56*). Moreover, strategies combining the avidity of scFvs and the targeting of two molecules (or epitopes) may represent interesting approaches to increase the antitumoral efficacy and to reduce the toxicity (*57, 58*). This would be particularly relevant for MM where, despite impressive initial responses, a significant number of patients experience a relapse after CAR T cell therapies (*5, 6*). However, the impact that CAR density could play on these approaches using CAR T cells with strong avidity and/or targeting several molecules, would be an additional aspect that should be further analyzed.

A relevant question faced in this work was the development of specific methodology for proper identification of CAR^High^ T cells from transcriptomic data. We defined a molecular signature associated with increased CAR density, for both CD4^+^ and CD8^+^ CAR T cells, that showed a higher correlation with cell surface CAR levels that the RNA expression. The implications of these signatures were severalfold: 1) this signature was key to annotate cells and identify clusters enriched in CAR^High^ T cells; 2) in addition, these signatures allowed us to perform GRN analysis using single cell data and identify key regulons associated to exhaustion of CAR T cells, such as NR4A1 and MAF (*43*–*45*), that were differentially activated in CAR^High^ T cells, providing mechanistic insights of the regulatory pathways driving the differences between CAR T cells with different CAR density; 3) finally, the definition of the signature associated with CAR^High^ T cells allowed us to apply the gene signature to transcriptomic data from CAR T cell products of patients undergoing CAR T treatment (*25, 26*), demonstrating a correlation between CAR expression and response in several cohorts of patients with different diseases (CLL and DLBCL). In the case of MM patients undergoing CAR T therapies, we also found a correlation between the CAR levels present on the therapeutic products and both the depth of response and its duration. These observations support our hypothesis that the presence of a high number of CAR^High^ T cells within the products limits the efficacy of CAR T therapies and suggests that the application of our gene signature may represent a new prognostic factor in patients treated with CAR T cells.

In summary, our data demonstrate that CAR density plays an important role in CAR T activity with an impact on clinical response. Moreover, the comprehension of regulatory mechanisms driven by CAR densities at the single cell level offers an important tool for the identification of key regulatory factors, that could be modulated for the development of improved therapies. Lastly, the application of a gene signature associated to increased CAR density, if validated in additional cohorts may represent a valuable tool for predicting responses in patients undergoing CAR T cell therapy.

## MATERIALS AND METHODS

### Cell lines

Jurkat-TPR (kindly provided by Dr. P. Steinberg; Medical University of Vienna), ARP-1-GFPLuc (kindly provided by Dr. Epstein; University of Arkansas for Medical Sciences), U266, MOLP8, K562 and K562-CD19 cells were cultured in RPMI 1640 supplemented with 10% FBS. MOLM13 and MV411 were cultured in RPMI 1640 supplemented with 20% FBS. HEK293T and BT474 cells were cultured in DMEM supplemented with 10% FBS. All media were supplemented with 1% penicillin/streptomycin and 1% L-Glutamine. All cell lines were maintained at 37°C in 5% CO_2_.

### Lentiviral vector construction and virus preparation

A third-generation self-inactivating lentiviral vector (pCCL) was used to express under the EF1a promoter a second-generation CAR constructs targeting BCMA (J22.9 clone from ARI-0002h), CD19 (FMC63 clone), CD33 (my96 clone) or HER2 (FRP5 clone). CAR structure comprised the single-chain variable fragment (scFv), a CD8 hinge and transmembrane domain, the 4-1BB and CD3ζ endodomains fused to a truncated version of the EGFR or a BFP reporter gene. Lentiviral vectors were produced in HEK293T cells following standard procedures. Briefly, 6×10^6^ cells were co-transfected with LV vector along with pMDLg/pRRE (Gag/Pol), pRSVRev and pMD2.G (VSVG envelope) packaging plasmid using Lipofectamine 2000 (Invitrogen). Supernatants were collected 40h after transfection, filtered, concentrated using Lenti-X Concentrator (Takara) following manufacturer specifications and stored at -80°C until use.

### Analysis of CAR signaling in Jurkat-TPR

Jurkat-TPR cells, transduced at MOI of 1 with CARs targeting BCMA, CD33, CD19 or HER2, were co-cultured in triplicate with ARP1-GFPLuc, MOLP8 and U266 cells for BCMA CARs, MOLM13 and MV4-11 cells for CD33, K562-CD19 cells for CD19 CARs or BT474 cells for HER2 CARs, at a 1:1 effector to tumor cell ratio. Non-transduced Jurkat-TPR cells were used as control. CAR^High^ and CAR^Low^ subpopulations were defined according to the fluorescence intensity (FI) using an EGFR-APC antibody (clone AY13, Biolegend). Thresholds for CAR^High^ and CAR^Low^ were define as the top and bottom FI quartiles respectively (CAR^High^: FI > 1.5×10^5^ and CAR^Low^: FI < 2×10^4^). Activation of the NFAT, NF-κB and AP-1 pathways was quantified before and 24h after co-culture with tumor cells measuring eGFP, eCFP and mCherry emissions respectively, using a CytoFLEX LX Flow Cytometer (Beckman Coulter) (Fig. S1).

### CAR T cell generation

CD4^+^ and CD8^+^ cells were isolated from PBMCs using CD4 and CD8 MicroBeads (Miltenyi Biotec) in the AutoMACS Pro Separator (Miltenyi Biotec). Isolated T cells were activated with 10 μl/ml T cell TransAct (Miltenyi Biotec) for 48h and infected with the CAR lentiviral vector at MOI 2 with 10 μl/ml of LentiBoost (Sirion Biotech). CAR T cells were expanded in RPMI 1640 culture medium supplemented with 3% human serum (Sigma), 1% penicillin/streptomycin and 625 IU/ml of human IL-7 and 85 IU/ml of human IL-15 (Miltenyi Biotec). CAR T cells were counted, and the concentration was adjusted to 1×10^6^ cells/ml every two days.

### Flow cytometry and CAR^High^ and CAR^Low^ T cell isolation

Phenotypic characterization of T cells and CAR T cells was performed at day 0 and 13 of the production, respectively. All antibodies were purchased from Biolegend unless otherwise stated (Table S5). Data was acquired on a BD FACSCanto II (BD Biosciences) and analyzed using the FlowJo Software version 10 (Tree Star). CAR^High^ and CAR^Low^ T cell subpopulations were sorted according to the BFP FI. Thresholds for CAR^High^ and CAR^Low^ were define as the top and bottom FI quartiles respectively (CAR^High^: FI > 1.2×10^4^; CAR^Low^: FI < 4×10^3^) (Fig. S3). All cells were sorted using a MoFlo Astrios EQ (Beckman Coulter).

### Quantification of CAR density

The number of CAR molecules on the surface of CAR T cell products from healthy donors and MM patients was quantitated using Quantum Simply Cellular (QSC) (Bangs Laboratories, USA: 815), according to the manufacturer’s protocol. This methodology allows the conversion of fluorescence intensity value into absolute numbers of binding molecules using a calibration curve. Data acquisition was performed on a BD FACSCanto II (BD Biosciences), and the results were analyzed using FlowJo Software version 10 (Tree Star). A threshold of 5000 and 1500 molecules was applied to define cells as CAR^High^ or CAR^Low^ respectively, since these were a lower bound (5000 on CAR^High^) and an upper bound (1500 on CAR^Low^) on number of molecules present on the samples sorted for functional and transcriptomic experiments.

### Viral copy number

Viral copy number (VCN) per cell were determined by qPCR. Genomic DNA was extracted using the DNeasy Blood and Tissue Kit (Qiagen). VCN/cell were quantified by duplex detection of the Psi sequence, normalized to ALBUMIN, using specific primers and detected with the TaqMan probes (Table S6). qPCR was performed using the Absolute qPCR Mix Low ROX Mix (Thermo Scientific) in a QuantStudio™ 3 Real-Time PCR System (Thermo Fisher Scientific). Results were analyzed in QuantStudio 3 Design and Analysis Software (Thermo Fisher Scientific).

### Cytotoxicity assay and cytokine production

Cytotoxicity was determined using ARP1-GFPLuc as target tumor cells. Briefly, ARP1-GFPLuc cells were cultured with CAR^High^ T and CAR^Low^ T cells at different ratios in RPMI 1640 culture medium, supplemented with 3% human serum (Sigma) and 1% penicillin/streptomycin in Nunc™ 96-Well Round Bottom plates (ThermoFisher Scientific). After 24h, luminescence measured using the Bright-Glo™ Luciferase Assay System (Promega) according to the manufacturer’s instructions. IFNγ, TNFα and IL-2 cytokine production was quantified using BD™ Immunoassay ELISA reagents (BD Biosciences) following manufacturer protocol.

### *In vivo* experiments

All experimental procedures were approved by the Ethics Committee of the University of Navarra and the Institute of Public Health of Navarra according to European Council Guidelines. NOD-SCID-Il2rg^−/−^ (NSG) mice were purchased from The Jackson Laboratory (JAX) and bred and maintained in-house in a pathogen-free facility. Eight-to-twelve-week-old male or female mice were irradiated at 1.5 Gy at day -1 and 1 × 10^6^ ARP1-GFPLuc cells were intravenously injected the following day. Mice were randomized to ensure equal pre-treatment tumor burden before CAR T cell treatment. At day 6 mice received i.v. injection of 0.5×10^6^ either CAR^High^ T or CAR^Low^ T cells. A subset of mice was sacrificed at day 28 to analyze the presence of cells in bone marrow. Tumor progression was measured by bioluminescent imaging using the PhotonIMAGER (Biospacelab). Signal was quantified using M3Vision Analysis Software (Biospacelab). Mice were humanely euthanized when mice demonstrated signs of morbidity and/or hindlimb paralysis.

### RNA-sequencing and bioinformatics analysis

RNA-seq was performed following MARS-seq protocol adapted for bulk RNA-seq (*59, 60*) with minor modifications. RNA-seq libraries quantification was done with Qubit 3.0 Fluorometer (Life Technologies) and size profiles were examined using Agilent’s 4200 TapeStation System. Libraries were sequenced in an Illumina NextSeq500 at a sequence depth of 10 million reads per sample. Samples were aligned to the Human genome (GRCh38) with STAR (v2.6.1). Gene expression was quantified with quant3p (github.com/ctlab/quant3p). Downstream analyses were performed in R (v3.6.2). Data transformation, normalization, and differential gene expression analysis were performed with DESeq2. T cell activation and tonic signaling gene signatures used in this work were obtained from previous publications (*25*–*28*) (Table S7).

### Assay for transposase-accessible chromatin (ATAC-seq) and bioinformatics analysis

Accessible chromatin mapping was performed using FAST-ATAC-seq (*61*) with minor modifications. Libraries were quantified and their size profiles examined as described above. Sequencing was carried out in an Illumina NextSeq500 at a depth of 20 million reads per sample. ATAC-seq reads were aligned to the human genome (GRCh38) using Bowtie2 (v2.3.4). Peak calling from each individual replicate was performed with MACS2 (v.2.1.0). Differential accessible analysis was analyzed with Csaw R package. The consensus peak set was derived from the union of all replicate peak sets for both conditions. For normalization, a non-linear LOESS-based (LOESS: Locally estimated scatterplot smoothing) method was applied. Differential enrichment was analyzed using edgeR package. Finally, ChIPseeker R package was used for annotation and visualization of genomics features.

### Single cell RNA-sequencing (scRNA-seq)

scRNA-seq was performed in FACS-sorted CAR T cells (BFP^+^ cells) from three independent donors using the Chromium Single Cell 5′ Reagent Kit (10X Genomics) according to the manufacturer’s instructions. After quality control and quantification, single cell libraries were sequenced at an average depth of at least 30000 reads/cell. 43,981 cells were analyzed, and 28,117 cells passed quality control with an average sequencing depth of 42,296 reads/cell, yielding an average of 2,541 genes/cell. TCR α/β sequencing was performed with 10X Genomics Single Cell V(D)J Immune Profiling Solution (10× Genomics). After quality control and quantification, single cell V(D)J enriched libraries were pooled and sequenced at a minimum depth of 5000 reads per cell.

scRNA-seq data were demultiplexed, aligned to the human reference (GRCh38) and the feature-barcode matrix was quantified using Cell Ranger (v6.0.1) from 10X Genomics. Further computational analysis was performed using Seurat (v3.1.5). Cells were subjected to QC filters based on the number of detected genes, number of UMIs and proportion of UMIs mapped to mitochondrial and ribosomal genes per cell. Each dataset was subjected to normalization, identification of highly variable genes and removal of unwanted sources of variation. Integration of all the dataset was based on Seurat’s canonical correlation analysis. Unsupervised clustering analysis with the resolution set to 0.8 yield a total of 23 cell clusters. Non-linear dimensional reduction was performed using t-distributed stochastic neighbor embedding (t-SNE) and UMAP. To describe the cell types and states defined by each cluster, we performed a manual review of the differentially expressed genes that were identified for each cell cluster by Seurat, using canonical marker genes as reference. TCR reconstruction and paired TCR clonotype analysis was performed using Cell Ranger v6.0.2 for V(D)J sequence assembly.

### Generation of CAR^High^ signature

The CAR^High^ signature was generated using differentially expressed genes between CAR^High^ T and CAR^Low^ T cell samples for each cell type (CD4^+^ and CD8^+^ cells). Specifically, the signature is created as:

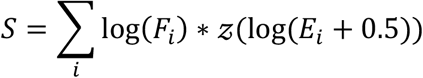

where *F*_*i*_ is the fold change of the gene *i* between the CAR^High^ T and CAR^Low^ T samples, 𝒵(.) is the function computing the z-score and *E*_*i*_ is the normalized gene expression for the *i*-th gene. This signature takes the z-score of the logarithm of the expression gene, allowing to compare the genes between them, and multiplying it by the logarithm of the fold-change to imprint the directionality of the signature gene.

### Gene regulatory network analysis

Cells were labelled as CAR^High^ by applying the developed CAR^High^ signature. Then, using the most variable 300 TFs and 3000 genes, SimiC was run with the default parameters across CAR^High^-labelled cells and the rest. GRNs were plotted using the GRN incidence matrices provided by SimiC. The histograms for the different regulons were computed from the ‘Regulon Activity Score’ provided by SimiC. This score was also used to compute the regulatory dissimilarity score for the selected cell clusters.

### Patient samples and clinical data

Samples were obtained from patients with MM enrolled in the academic clinical trial CARTBCMA-HCB-01 (NCT04309981), lead by Dr. Fernandez de Larrea (Hospital Clínic in Barcelona), assessing the BCMA-CAR T ARI-0002h, developed at IDIBAPS/Hospital Clínic in Barcelona. All subjects provided written informed consent. Clinical response was defined according to the MM Response Criteria (*62*) as: Stringent Complete Response (sCR), Very Good Partial Response (VGPR), Partial Response (PR) and Progressive Disease (PD). The CAR^High^ signature was applied to gene expression data publicly available (*25, 26*), which allowed us to: i) quantify the number of CAR^High^ T cells within CD4^+^ and CD8^+^ population in CAR T cell products when scRNA-seq data was available; and ii) to score the CAR T cell products according to CD4^+^ and CD8^+^ CAR^High^ signature when bulk RNA-seq data was available. The clinical data was judged the same way as in the original manuscript, grouping the different treatment responses into: Non Response (NR), Partial Response (PR), Partial Response with Transformed Disease (PRTD) and Complete Response (CR).

### Statistical Analysis

Statistical analyses were performed using GraphPad Prism for Mac version 9.3.1. The different tests used in this work are indicated in the figure legend.

## Data Availability

All data produced in the present study are available upon reasonable request to the authors

## Acknowledgments

We thank the members of Hematology and Cell Therapy Department of the Clinica Universidad de Navarra for input throughout the course of the project and all the patients as well as families who made this study possible. We particularly acknowledge the patients for their participation in the Clinical trial CARTBCMA-HCB-01 (NCT04309981) and the Biobank of the University of Navarra for its collaboration.

## Author contributions

P.R-M., M.E.C-C., G.S, M.H., J.R.R-M., and F.P. designed the experiments. P.R-M., M.E.C-C., G.S, A.M-M. and C.C. conducted the experiments. P.R-M., M.E.C-C., G.S, A.O-C., M.H. and J.R.R-M. performed the data analysis. M.L.P-B., P.R-O., A.A. and J.S-M. provided clinical advice. A.O-C., M.E-R., C.C., M.C.V., M.R. M.P., M.J., A.U., C.F-L. and B.P. provided clinical samples and data. T.L., J.J.L., B.P., M.H., J.R.R-M., and F.P discussed the study design and the results. P.S-M., A.V-Z., S.R-D., R.M-T. and D.A. provided technical assistance. M.H., J.R.R-M., and F.P. were responsible for research supervision, coordination, and strategy. P.R-M. and J.R.R-M. drafted the manuscript. J.J.L., J.S-M., C.F-L., M.H., J.R.R-M., and F.P. reviewed and edited the manuscript. All authors reviewed and approved the final version of the manuscript.

## Data and materials availability

All data needed to evaluate the conclusions in the paper are present in the paper and/or the Supplementary Materials. The RNA-seq, ATAC-seq, and scRNA-seq data generated in this study have been deposited in the GEO database (GEO#) and will be available upon publication. Additional data related to this paper may be requested from the authors.

## Supplementary Materials

Supplementary material for this article includes:

Extended Material and Methods

Fig. S1. CAR density influences CAR-mediated signaling.

Fig. S2. *In vitro* characterization of CAR T cells targeting BCMA.

Fig. S3. Characterization of CAR T cells with different CAR densities.

Fig. S4. Phenotypic characterization of CAR T cells with different CAR densities.

Fig. S5. Epigenetic characterization of CAR^High^ T and CAR^Low^ T cells.

Fig. S6. Transcriptomic profile and chromatin landscape of CD4^+^ CAR^High^ T cells.

Fig. S7. Characterization of CAR T cells at single cell level.

Fig. S8. Generation and characterization of gene signatures associated to CAR^High^ T cells.

Fig. S9. Characterization of CAR^High^ T cells at single cell level.

Fig. S10. Analysis of GRNs in cell with different CAR densities.

Table S1. List of differentially expressed genes in RNA-seq analysis between CAR^High^ T and CAR^Low^ T cells in both CD4^+^ and CD8^+^ cell subsets.

Table S2. List of differential peaks in ATAC-seq analysis between CAR^High^ T and CAR^Low^ T cells in both CD4^+^ and CD8^+^ cell subsets.

Table S3. List of VDJ clonotypes detected in scRNA-seq analysis of CAR T cells.

Table S4. List of the genes used for the generation of the signatures for CD4^+^ and CD8^+^ CAR^High^ T cells.

Table S5. List of the antibodies used in this study.

Table S6. List of the primer and probe sequences used in this study.

Table S7. List of the genes used in the T cell activation and tonic signaling signatures.

