## Supplementary Materials for "CAR Density Influences Antitumoral Efficacy of BCMA CAR T cells and Correlates with Clinical Outcome"

**This PDF file includes:**

Extended Material and Methods
Figs. S1 to S10
Legends for tables S1 to S7

**Other Supplementary Materials for this manuscript include the following:**

Tables S1 to S7

### Extended Material and Methods

#### Lentiviral vector production.

Lentiviral vectors were produced in HEK293T cells following standard procedures. Briefly, $6\times10^6$ cells were plated and co-transfected 24h later with 6,9µg of LV vector along with 3,41µg of pMDLg/pRRE (Gag/Pol), 1,7µg of pRSVRev and 2µg of pMD2.G (VSVG envelope) packaging plasmid using Lipofectamine 2000 (Invitrogen). Supernatants were collected 40h after transfection and filtered with low retention 0.45 µm filters. Viral particles were concentrated using Lenti-X Concentrator (Takara) following manufacturer specifications and store at -80°C until use.

#### CAR T cell generation.

PBMCs were isolated from peripheral blood of healthy donors using Ficoll-Paque™ Plus (GE Healthcare) and then $CD4^+$ and $CD8^+$ cells were isolated using CD4 and CD8 MicroBeads (Miltenyi Biotec) in the AutoMACS Pro Separator (Miltenyi Biotec). Isolated T cells were cultured at a concentration of $1\times10^6$ cells/ml in RPMI 1640 culture medium (Lonza) supplemented with 3% human serum (Sigma), 1% penicillin/streptomycin (Gibco) and 625 IU/ml of human IL-7 and 85 IU/ml of human IL-15 (Miltenyi Biotec), and activated with 10 µl/ml T cell TransAct (Miltenyi Biotec). After 48h, T cells were infected with the CAR lentiviral vector at MOI 2 and 10 µl/ml of LentiBoost (Sirion Biotech), to enhance viral infection. CAR T cells were expanded in RPMI 1640 culture medium (Lonza) supplemented with 3% human serum (Sigma), 1% penicillin/streptomycin (Gibco) and 625 IU/ml of human IL-7 and 85 IU/ml of human IL-15 (Miltenyi Biotec). CAR T cells were counted, and the concentration was adjusted to $1\times10^6$ cells/ml every two days until day 13-14 of the production.

#### Viral copy number.

Viral copy number (VCN) per cell were determined by qPCR. Genomic DNA was extracted using the DNeasy Blood and Tissue Kit (Qiagen). VCN/cell were quantified by duplex detection of the Psi sequence, normalized to ALBUMIN, using specific primers and detected with the TaqMan probes. Primers and probes sequences are specified in Table S6. qPCR was performed in 12,5 µl final volume with 25 ng of gDNA using the Absolute qPCR Mix Low ROX Mix (Thermo Scientific) with 10 µmol of primers and TaqMan probes, in a QuantStudio™ 3 Real-Time PCR System (Thermo Fisher Scientific) with the following conditions: 95°C 10 min; 40 cycles of 95°C 15s, 60°C 1min; 95 °C 10 min. Results were analyzed in QuantStudio 3 Design and Analysis Software (Thermo Fisher Scientific).

#### RNA-sequencing and bioinformatics analysis.

RNA-seq was performed following MARS-seq protocol adapted for bulk RNA-seq with minor modifications. Briefly, 7000 to 10000 cells were sorted in 100µl of Lysis/Binding Buffer (Ambion), vortexed and stored at -80ºC until further processing. Poly-A RNA was reverse-transcribed using poly-dT oligos carrying a 7 nt-index. Pooled samples were subjected to linear amplification by IVT. Resulting aRNA was fragmented and dephosphorylated. Ligation of partial Illumina adaptor sequences was followed by a second reverse-transcription reaction. Full Illumina adaptor sequences were added during final library amplification. RNA-seq libraries quantification was done with Qubit 3.0 Fluorometer (Life Technologies) and size profiles were examined using Agilent's 4200 TapeStation System. Libraries were sequenced in an Illumina NextSeq500 at a sequence depth of 10 million reads per sample. Samples were demultiplexed using Illumina bcl2fastqsoftware (v1.2.4) and aligned to the Human (GRCh38) genome with STAR (v2.6.1). Sequencing data quality was assessed with FastQC software and Illumina adapter sequences, polyA tails, and short reads (less than 20 bases) were trimmed using Cutadapt (v1.18). Gene expression was quantified with quant3p (github.com/ctlab/quant3p). Downstream analyses were performed in R (v3.6.2). Minimal gene expression was established at 5cpm, and genes with lower levels of expression in more than 20% of samples from at least one sample group were removed for increased consistency. Data transformation, normalization, and differential gene expression analysis were performed with DESeq2. Data was processed using *RemoveBatchEffect* function from Limma analysis package for data visualization purposes.

**Assay for transposase-accessible chromatin (ATAC-seq) and bioinformatics analysis.** Accessible chromatin mapping was performed using FAST-ATAC-seq with minor modifications. Briefly, 10000 $CD4^+$ or $CD8^+$ $CAR^{High}$ T or $CAR^{Low}$ T cells from six independent donors were sorted in 1X PBS, 0.05% BSA and pelleted by centrifugation at 500 rcf for 5 min at 4 °C with low acceleration and brake settings in a pre-cooled swinging-bucket rotor centrifuge. Next, 25 µL of transposase mix (15 µL of 2x TD buffer (Illumina); 1 µL of TDE1 (Illumina), 0.25 µL of 5% digitonin (Promega); 8.75 µL of nuclease- free water) were added to the cells and the pellet was disrupted by pipetting. Transposition reactions were incubated at 37 °C for 30 min in an Eppendorf ThermoMixer with shaking at 450 rpm. Reactions were stopped at 4 °C for 5 min. To release tagmented DNA, samples were incubated at 40 °C for 30 min with 5 µL of clean up buffer (900 mM NaCl (Sigma), 30 mM EDTA (Millipore), 2 µL of 5% SDS (Millipore) and 2 µL of Proteinase K (NEB)). DNA was purified using a 2X SPRI beads cleanup kit (Agencourt AMPure XP, Beckman Coulter). To determine the total number of PCR cycles needed for library amplification, two sequential PCRs were performed using KAPA HiFi DNA Polymerase and customized Nextera PCR indexing primers (IDT). The conditions of the first PCR were: 5 min at 72 °C and 2 min at 98 °C followed by 9 cycles of 20 secs at 9 °C, 30 secs at 63 °C, and 1 min at 72 °C. Depending on the library concentration in this first PCR, a second PCR (2 min at 98 °C followed by 4 to 6 cycles of 20 secs at 98 °C, 30 secs at 63 °C, and 1 min at 72 °C) was performed aiming for a library concentration in the range of 2 to 10 ng/µL. PCR products were purified using a 2X SPRI beads cleanup. Libraries were quantified and their size profiles examined as described above. Sequencing was carried out in an Illumina NextSeq500 using paired-end, dual-index sequencing (Read1: 38 cycles; Read2: 38 cycles; i7 index: 8 cycles; i5 index: 8 cycles) at a depth of 20 million reads per sample. ATAC-seq reads were analyzed for quality control measures prior alignment to the human genome assembly (GRCh38) using Bowtie2 (v2.3.4). For each sample, mitochondrial reads, ummapped reads and low mappability (< 30) reads were filtered out using samtools (v1.9). Library complexity and removal of PCR duplicates were done with Picard tools (v2.18.17). Reads in blacklisted genomic regions as defined by ENCODE were removed. All downstream analyses were performed on these filtered reads. Peak calling from each individual replicate was performed with MACS2 (v.2.1.0). Differential accessible analysis was analyzed with Csaw R package. The consensus peak set was derived from the union of all replicate peak sets for both conditions. Then, reads were count in the specified consensus peak set with *regionCounts* function and low abundance peaks were subsequently filtered (logCPM > -3 threshold). For normalization, a non-linear LOESS-based (LOESS: Locally estimated scatterplot smoothing) method was applied. Differential enrichment was analyzed using edgeR package and *getBestTest* function was used to get the most significant window as a statistical representation. The final window was filtered with significant changes being defined by FDR value<0.05, adj. p<0.05 and absolute FC>= 1.4 (gain or loss). Finally, ChIPseeker R package was used for annotation and visualization of genomics features.

**Single cell RNA sequencing (scRNA-seq)**
FACS-sorted CAR T cells ($BFP^+$ cells) from three independent donors were interrogated using the 10X Chromium platform for simultaneous transcriptome and TCR profiling.

*5' Gene expression Profiling.* scRNA-seq was performed using the Chromium Single Cell 5′ Reagent Kit (10X Genomics) according to the manufacturer's instructions. Briefly, CAR T cells were sorted in 100µl 1X PBS, 0.04% BSA and their concentration and viability were determined using Nexcelom's Cellometer K2 Fluorescent Cell Counter. Cell concentration was adjusted to 700 to 1200 cells/µl to maximize the likelihood of achieving the desired cell recovery target. Then single-cell suspension was mixed with RT Master Mix and loaded together with barcoded single-cell 5′ gel beads and partitioning oil into separate single cell A Chips to generate Gel Beads in Emulsion (GEMs) using Chromium Controller. Thus, individual cells were lysed and the polyadenylated RNA was reverse-transcribed and barcoded inside each GEM. Barcoded complementary DNA (cDNA) was recovered through post GEM-RT cleanup and PCR-amplified. cDNA was quantified with Qubit dsDNA HS Assay Kit (Thermo Fisher Scientific) and its profile examined using High Sensitivity D5000 ScreenTape Assay (Agilent Technologies). Fifty nanograms of double-stranded cDNA were used for 5′ Gene Expression library construction. cDNA was fragmented, end-repaired, A-tailed and ligated to an adaptor. A final PCR-amplification with barcoded primers was performed for sample indexing. Sequencing was performed in a NextSeq500 (Illumina) (Read1: 26 cycles; Read2: 57 cycles; i7 index: 8 cycles) at an average depth of at least 30000 reads/cell.

*Single-cell TCR repertoire sequencing.* TCR α/β sequencing was performed with 10X Genomics Single Cell V(D)J Immune Profiling Solution (10× Genomics). Briefly, full-length V, D and J gene segments were amplified from barcoded cDNA using a Chromium Single Cell V(D)J Enrichment Kit (human T cells) by means of two sequential PCRs with primers targeting the adaptor and the gene constant region. Enriched cDNA was quantified as described above and 50 ng were further used for library construction as described above. After quality control and quantification, sequencing of Single Cell V(D)J enriched libraries was externalized to Macrogen Inc. (http://www.macrogen.com/) at a minimum depth of 5000 reads per cell.

**scRNA-seq bioinformatics.**
*Raw sequencing data processing, QC, data filtering and normalization.* The raw scRNA-seq data were demultiplexed, aligned to the Human (GRCh38) reference and the feature-barcode matrix was quantified using Cell Ranger (v6.0.1) from 10X Genomics. Further computational analysis was performed using Seurat (v3.1.5). Cells were subjected to QC filters based on the number of detected genes, number of UMIs and proportion of UMIs mapped to mitochondrial and ribosomal genes per cell. The thresholds filters for each variable were selected based on their distribution within each of the single cell libraries and visual inspection of quality control scatter plots. Genes detected in fewer than three cells and cells where fewer than 150 genes had nonzero counts were filtered out and excluded from subsequent analysis. Low-quality cells where more than 10% and more than 40% mitochondrial or ribosomal content respectively, were also discarded. In addition, cells with more than the 97.5$^{th}$ percentile of detected genes per sample, were discarded to remove likely doublet or multiplet captures.

*Unsupervised cell clustering and dimensionality reduction.* Each dataset was subjected to normalization, identification of highly variable genes and removal of unwanted sources of variation using Seurat (v3.1.5). Integration of all the dataset was based on Seurat's canonical correlation analysis. Different resolution parameters for unsupervised clustering were examined to determine the optimal number of clusters. For this study, the first 40 PCs and 3000 highly variable genes identified by Seurat were used for sample integration and unsupervised clustering analysis with the resolution set to 0.8, yielding a total of 23 cell clusters. Non-linear dimensional reduction was performed using t-distributed stochastic neighbor embedding (t-SNE) and UMAP.

*Inferring cell cycle stage and DEGs.* The cell cycle stage was computationally assigned to each individual cell using the R code implemented in Seurat based on expression profiles of the cell-cycle-related signature genes Differential expressed genes (DEGs) were identified for each cluster using the *FindConservedMarkers* function of the Seurat R package with default settings. Heat maps were then generated using the *heatmap* function in pheatmap R package for filtered DEGs.

*Determination of major cell types and cell states.* The major cell type (CD4 and CD8) was defined by marker gene expression (*CD4*, *CD40LG*, *TNFRSF4*, *CD8A* and *CD8B*) by 10X transcriptome. To describe the cell types and states (activated, memory, exhausted, etc…) defined by each cluster, we performed a manual review of the DEGs that were identified for each cell cluster by Seurat, using canonical marker genes as reference.

*TCR V(D)J sequence assembly, paired clonotype calling, TCR diversity and clonality analysis.* TCR reconstruction and paired TCR clonotype analysis was performed using Cell Ranger v6.0.2 for V(D)J sequence assembly.

### Supplemental Figures

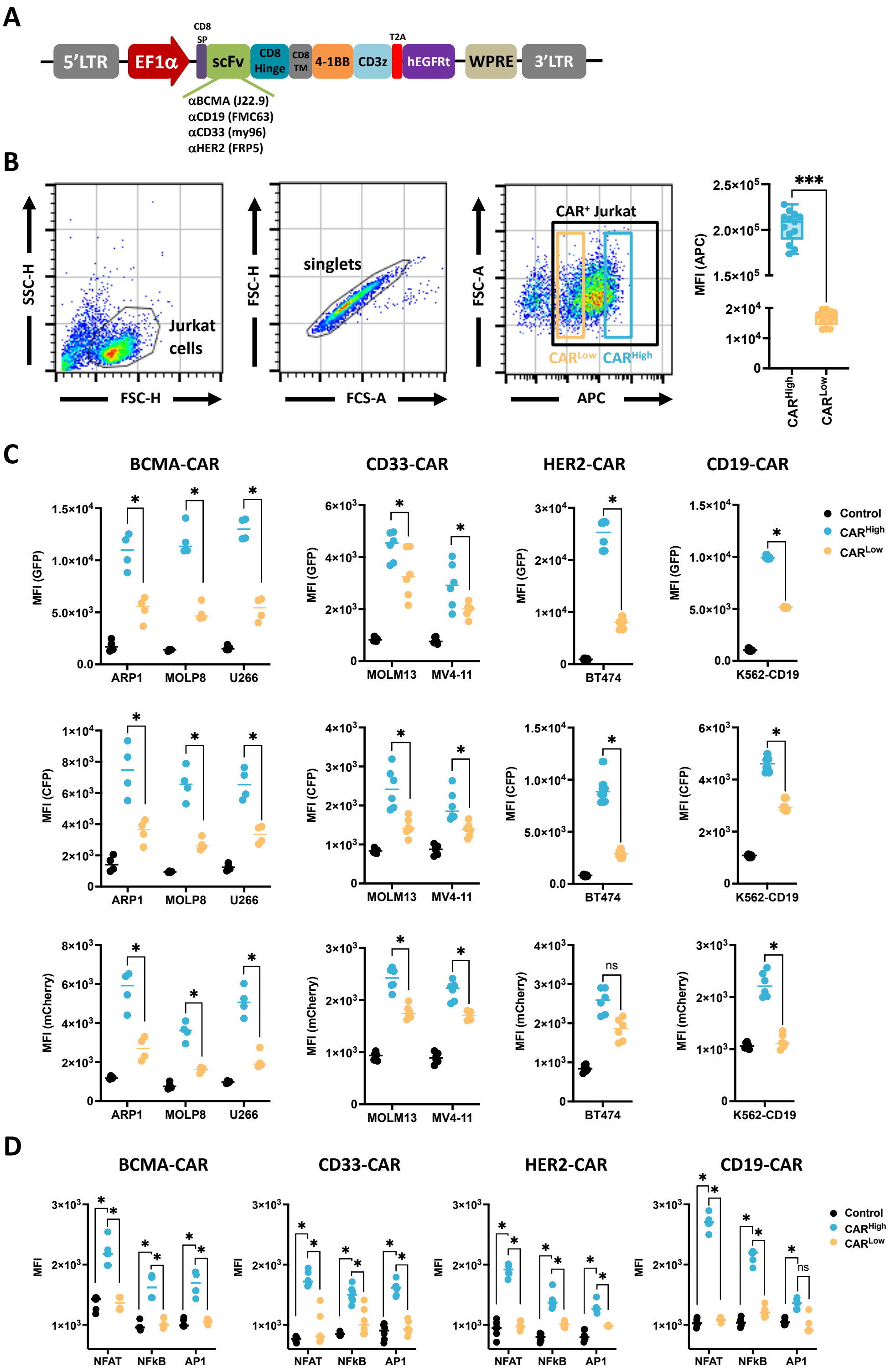


**Fig. S1. CAR density influences CAR-mediated signaling.** A triple parameter reporter (TPR) system in Jurkat cells was used to measure, by flow cytometry, CAR-mediated activation of the main signaling pathways (NFAT, NFkB and AP1) before and after tumor recognition. **(A)**

Schematic representation of the lentiviral vector used in these experiments to express a second-generation CAR construct under EF1a promoter. CAR construct comprises a scFv targeting either BCMA, CD33, HER2 or CD19, the transmembrane (TM) and hinge domains derived from CD8a, the intracellular 4-1BB costimulatory and CD3z signaling domains, fused, via T2A, to a truncated EGFR reporter sequence. **(B)** Gating strategy to define $CAR^{High}$ and $CAR^{Low}$ subpopulations according to the fluorescence intensity (FI) of EGFR-APC ($CAR^{High}$: FI > $1.5x10^5$; $CAR^{Low}$: FI < $2x10^4$). **(C)** Measurement of the mean fluorescence intensity (MFI) of the NFAT (GFP; upper panels), NF-κB (CFP; middle panels) and AP-1 (mCherry; lower panels) pathways in $CAR^{High}$ and $CAR^{Low}$ subpopulations, 24h after co-culture of Jurkat TPR cells infected with the different CAR constructs with the indicated tumor cells. Non-infected Jurkat TPR cells were used as control. **(D)** Measurement of the MFI of the NFAT, NF-κB and AP-1 pathways in $CAR^{High}$ and $CAR^{Low}$ subpopulations in Jurkat TPR cells infected with the different CAR constructs in the absence of antigen stimulation (tonic signal). Non-infected Jurkat TPR cells were used as control. Wilcoxon test for paired samples (B) and Kruskal-Wallis test (C and D). ns: not significant; *p<0.05; ***p>0.001.

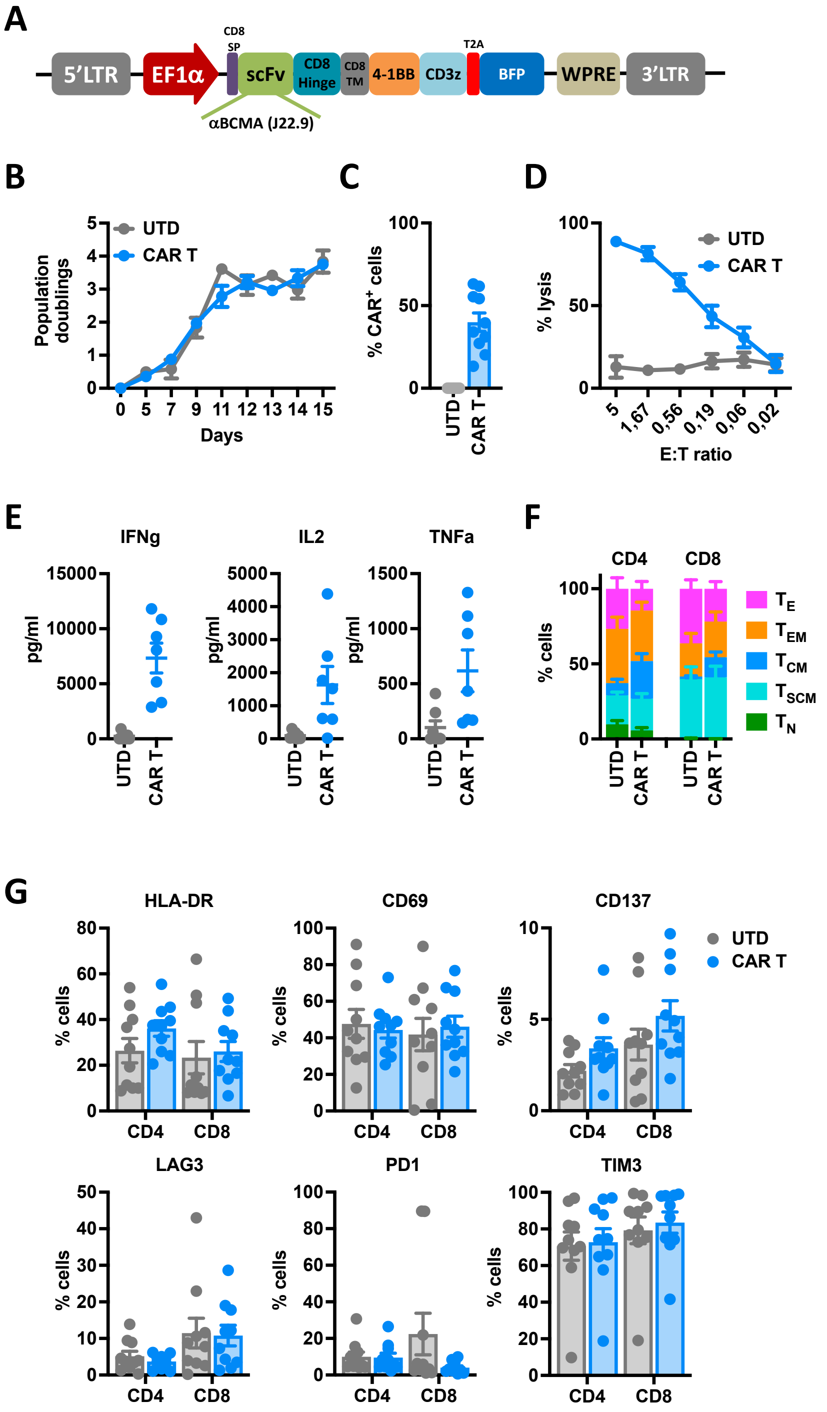


**Fig. S2. *In vitro* characterization of CAR T cells targeting BCMA.** CAR T cells from ten healthy donors that were generated and characterized. **(A)** Schematic representation of the lentiviral vector used for CAR T cell generation, expressing a second-generation CAR construct under EF1a promoter. CAR construct comprises a scFv targeting BCMA, the transmembrane (TM) and hinge domains derived from CD8a, the intracellular 4-1BB costimulatory and CD3z signaling domains, fused, via T2A, to a BFP reporter sequence. **(B)** Population doublings of the CAR T cells during the production (n=10). Non-transduced cells (UTD) from the same donor were used as control. **(C)** Percentage of transduced cells ($CAR^{+}$) at the end of each CAR T and UTD cell production (n=10).

**(D)** Quantification of the cytotoxic activity of CAR T and UTD cells against ARP1-GFPLuc at different E:T ratio. The percentage of specific lysis (average of three technical replicates) for each CAR T cell production (n=10) is depicted. **(E)** Quantification of IFNγ, IL-2 and TNFα levels in supernatants from cytotoxic assays (ratio 0.56:1) measured by ELISA. The cytokine concentration (pg/ml; average of three technical replicates) for each CAR T cell production (n=10) is depicted. **(F)** Analysis of the phenotype of CAR T and UTD cells at resting state for each T cell subpopulation. $T_N$: naïve; $T_{SCM}$: stem central memory; $T_{CM}$: central memory; $T_{EM}$: effector memory; $T_E$: effector. **(G)** Analysis of the expression of HLA-DR, CD69, CD137, LAG3, PD1 and TIM3 in CAR T and UTD cells at resting state. (B-G) Mean ± SEM of the ten independent productions is represented.

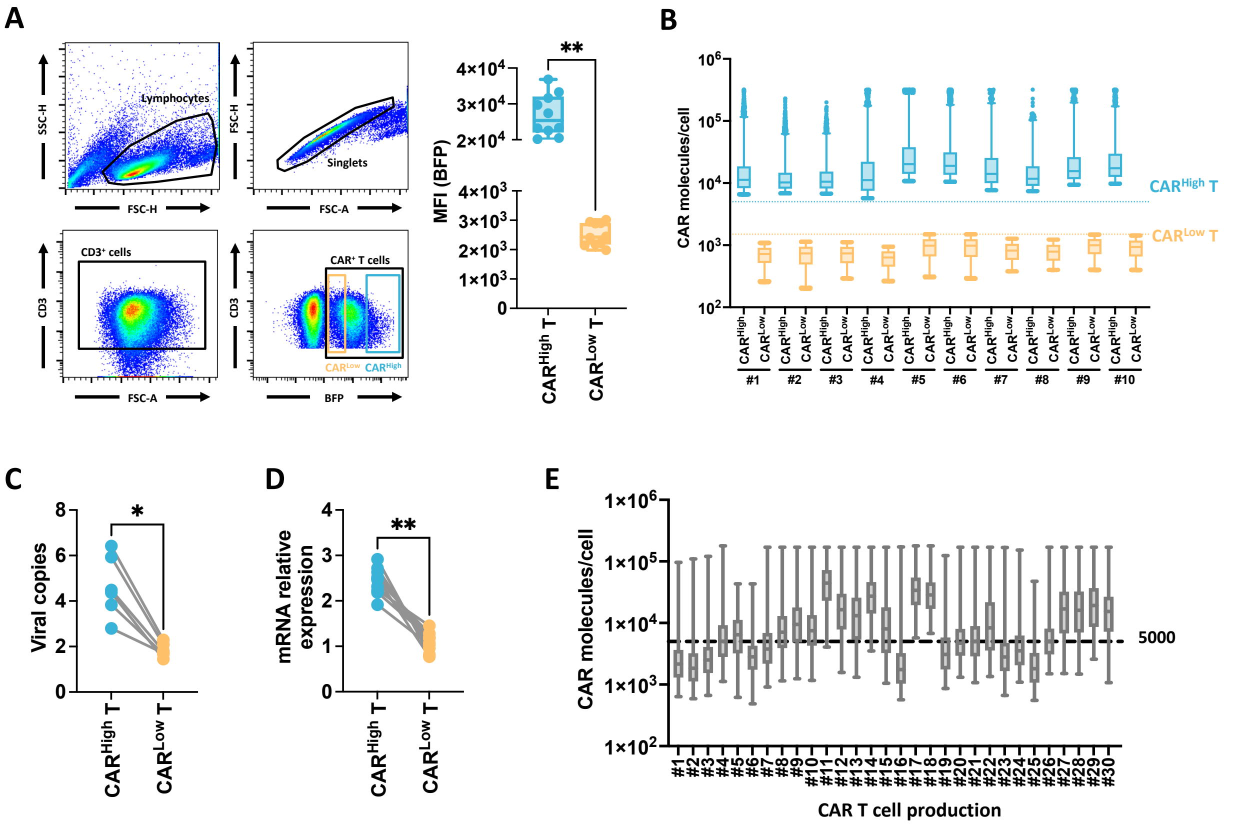


**Fig. S3. Characterization of CAR T cells with different CAR densities. (A)** Gating strategy to define CAR$^{High}$ T and CAR$^{Low}$ T subpopulations according to the fluorescence intensity (FI) of BFP. CAR$^{High}$ T cells include those cells with a BFP FI > 1.2x10$^4$ (average FI 26861 ± 5795), while CAR$^{Low}$ T subpopulation was restricted to BFP FI < 4x10$^3$ (average FI 2513 ± 388). These FI values corresponded to the top and bottom FI quartiles respectively. **(B)** Quantification of the number of CAR molecules on the surface of CAR$^{High}$ T and CAR$^{Low}$ T subpopulations for each CAR T cell production (n=10), using an antibody-binding capacity bead assay based on FI. CAR$^{High}$ T cells presented more than 5000 CAR molecules/cell, meanwhile CAR$^{Low}$ T cells were below 1500 molecules. Analysis of the copy number integrations of the LV vector **(C)** and CAR mRNA expression **(D)** in CAR$^{High}$ T and CAR$^{Low}$ T subpopulations isolated from 6 independent CAR T cells productions. **(E)** Quantification of the number of CAR molecules on the surface of the CAR T products from CARTBCMA-HCB-01 academic clinical trial (NCT04309981) using an antibody-binding capacity bead assay based on FI. The threshold of 5000 molecules/cell used to quantify the percentage of CAR$^{High}$ T cells is indicated. Wilcoxon test for paired samples (C and D). *p>0.05; **p<0.01.

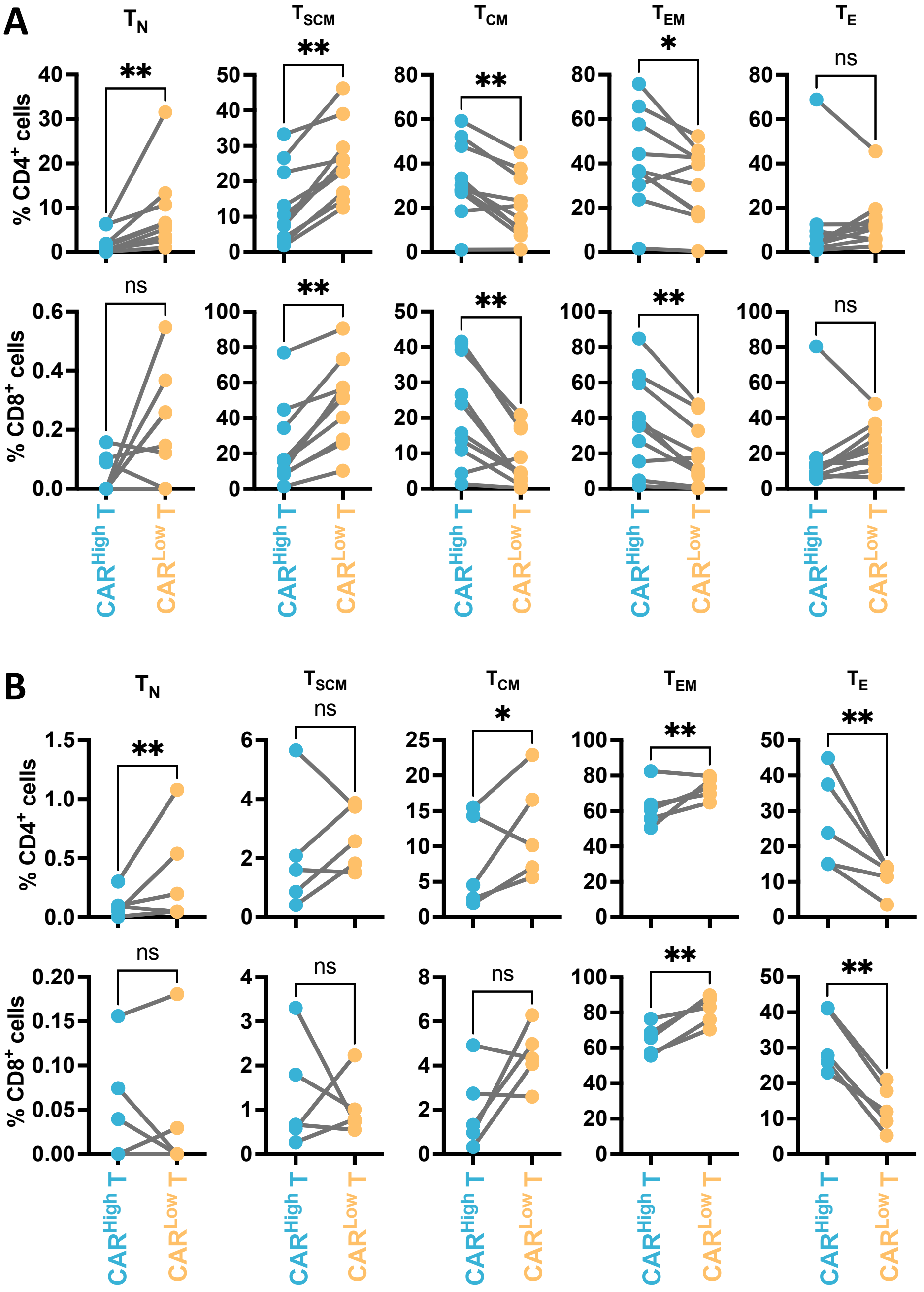


**Fig. S4. Phenotypic characterization of CAR T cells with different CAR densities.** Analysis of the phenotype of $CAR^{High}$ and $CAR^{Low}$ T cell populations before **(A)** (n=10) and after stimulation **(B)** (n=5) with ARP1-GFPLuc tumor cells in both, $CD4^+$ (upper line) and $CD8^+$ (Lower line) cell subset. Each T cell subpopulation is depicted. $T_N$: naïve; $T_{SCM}$: stem central memory; $T_{CM}$: central memory; $T_{EM}$: effector memory; $T_E$: effector. Wilcoxon test for paired samples (A and B). ns: not significant; *p<0.05; **p>0.01.

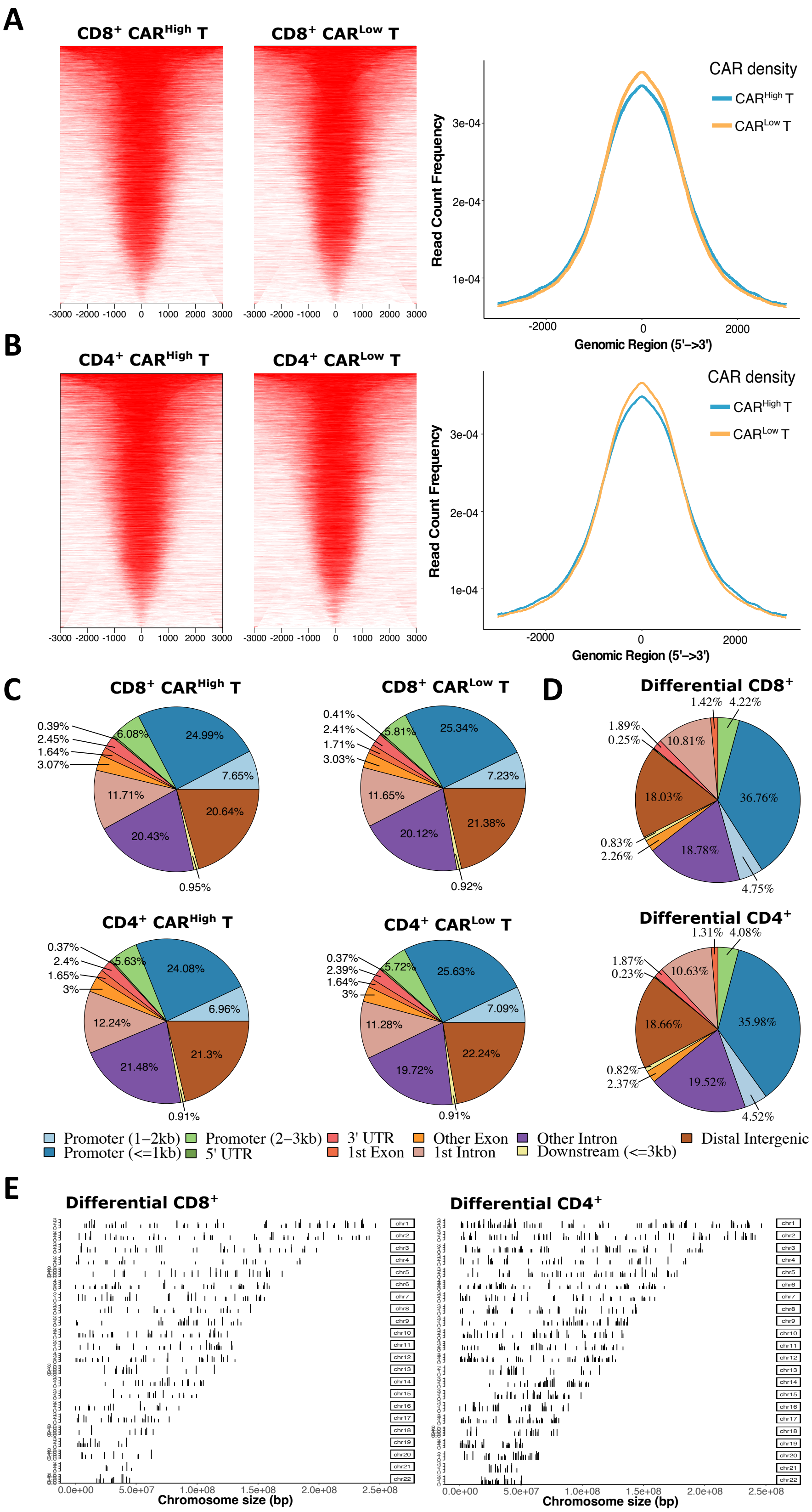


**Fig. S5. Epigenetic characterization of CAR$^{High}$ T and CAR$^{Low}$ T cells.** The epigenetic landscape of sorted CD4$^{+}$ and CD8$^{+}$ CAR$^{High}$ and CAR$^{Low}$ T cell populations (n=6) were profiled using assay for transposase-accessible chromatin (ATAC-seq). Heatmap (left) and quantification of read frequency (right) of ATAC-seq peaks related to TSS regions in $CD8^+$ **(A)** and $CD4^+$ **(B)** $CAR^{High}$ T and $CAR^{Low}$ T cell populations. **(C)** Pie plots showing the genomic annotation of ATAC-seq peaks in $CD8^+$ and $CD4^+$ $CAR^{High}$ T and $CAR^{Low}$ T cells. **(D)** Pie plots showing the genomic annotation of differential ATAC-seq peaks between $CAR^{High}$ T and $CAR^{Low}$ T cells in $CD8^+$ and $CD4^+$ cell subsets. **(E)** Plot showing the coverage of differential ATAC-seq peaks between $CAR^{High}$ T and $CAR^{Low}$ T cells over chromosomes in $CD8^+$ and $CD4^+$ cell subsets.

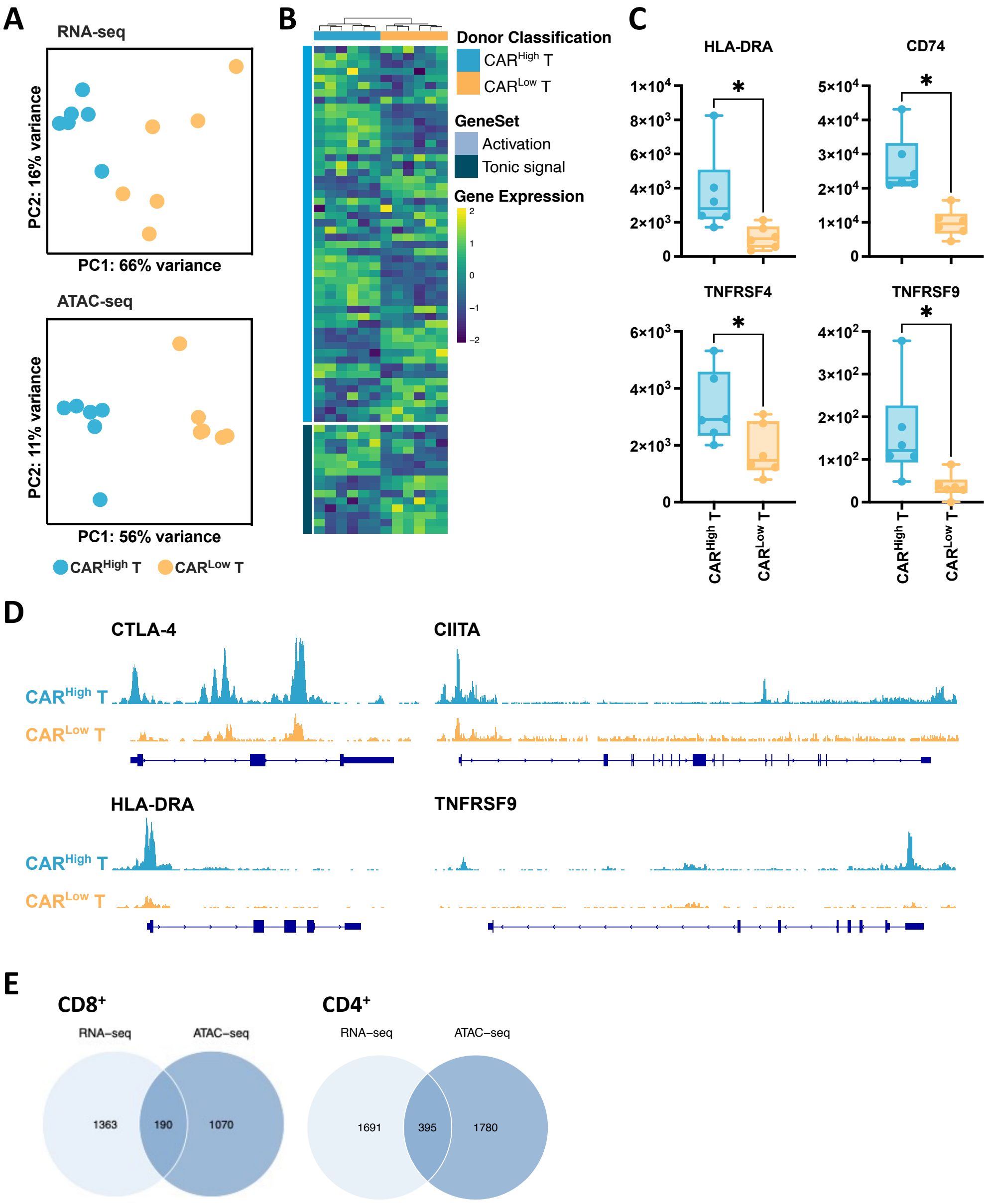


**Fig. S6. Transcriptomic profile and chromatin landscape of CD4$^{+}$ CAR$^{High}$ T cells.** The transcriptomic and epigenetic landscape of sorted CD4$^{+}$ CAR$^{High}$ and CAR$^{Low}$ T cells (n=6) were profiled using high-throughput RNA sequencing (RNA-seq) and assay for transposase-accessible chromatin (ATAC-seq). **(A)** RNA-seq and ATAC-seq principal components (PC) analysis, corrected by patient heterogeneity, of sorted CD4$^{+}$ CAR T cell subsets. Percentage of variance explained by PC1 and PC2 are depicted. **(B)** Heatmap of differentially expressed genes (DEGs) between CD4$^{+}$ CAR$^{High}$ T and CAR$^{Low}$ T cells associated to genes involved in tonic signaling and T cell activation. **(C)** Quantification of HLA-DR, CD74, TNFRSF4 (OX40) and TNFRSF9 (4-1BB) gene expression in CD4$^{+}$ CAR$^{High}$ T and CAR$^{Low}$ T cells. **(D)** UCSC genome browser tracks of CTLA-4, HLA-DRA, CIITA and TNFRSF9 showing differential peaks from ATAC-seq analysis between CD4$^{+}$ CAR$^{High}$ T and CAR$^{Low}$ T cells. (E) Venn diagram illustrating the overlapping between DEGs and differential peaks observed after the analysis of CAR$^{High}$ T and CAR$^{Low}$ T cells in CD8$^{+}$ and CD4$^{+}$ cell subsets. **(E)** Wilcoxon test for paired samples (C). *$p<0.05$.

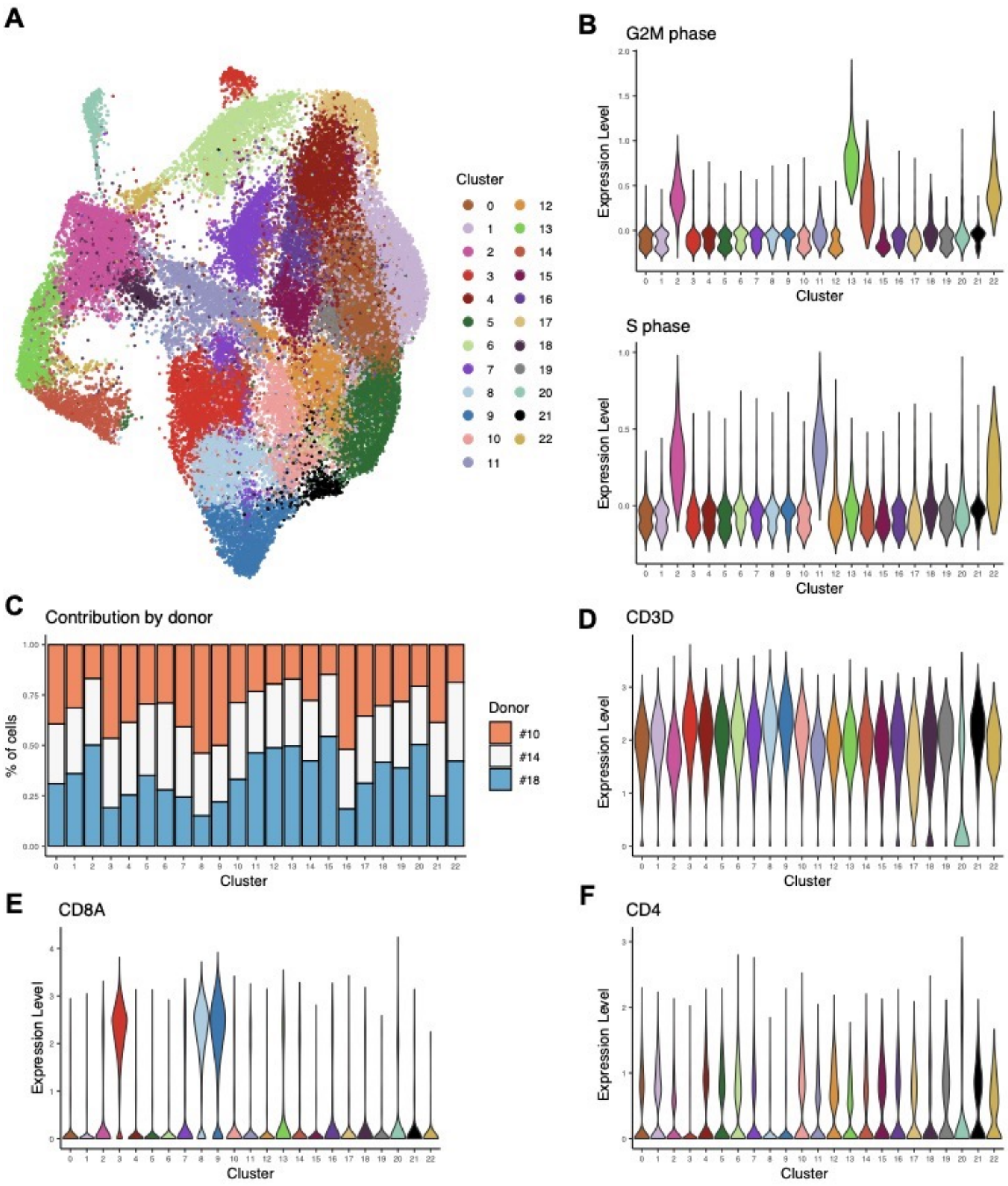


**Fig. S7. Characterization of CAR T cells at single cell level.** CAR+ (BFP+) cells from three independent CAR T cell productions were assayed by single-cell RNA sequencing. **(A)** An overview of the 43,981 cells analyzed in this study. UMAP plot showing the 23 clusters that were identified. **(B)** Violin plots showing the degree of G2M phase (upper panel) and S phase (lowe panel) signatures in each cluster. **(C)** Bar plot showing the percent contribution of cell from each donor to the different clusters. Violin plots showing the expression levels of CD3D **(D)**, CD8A **(E)** and CD4 in each cluster **(F)**.

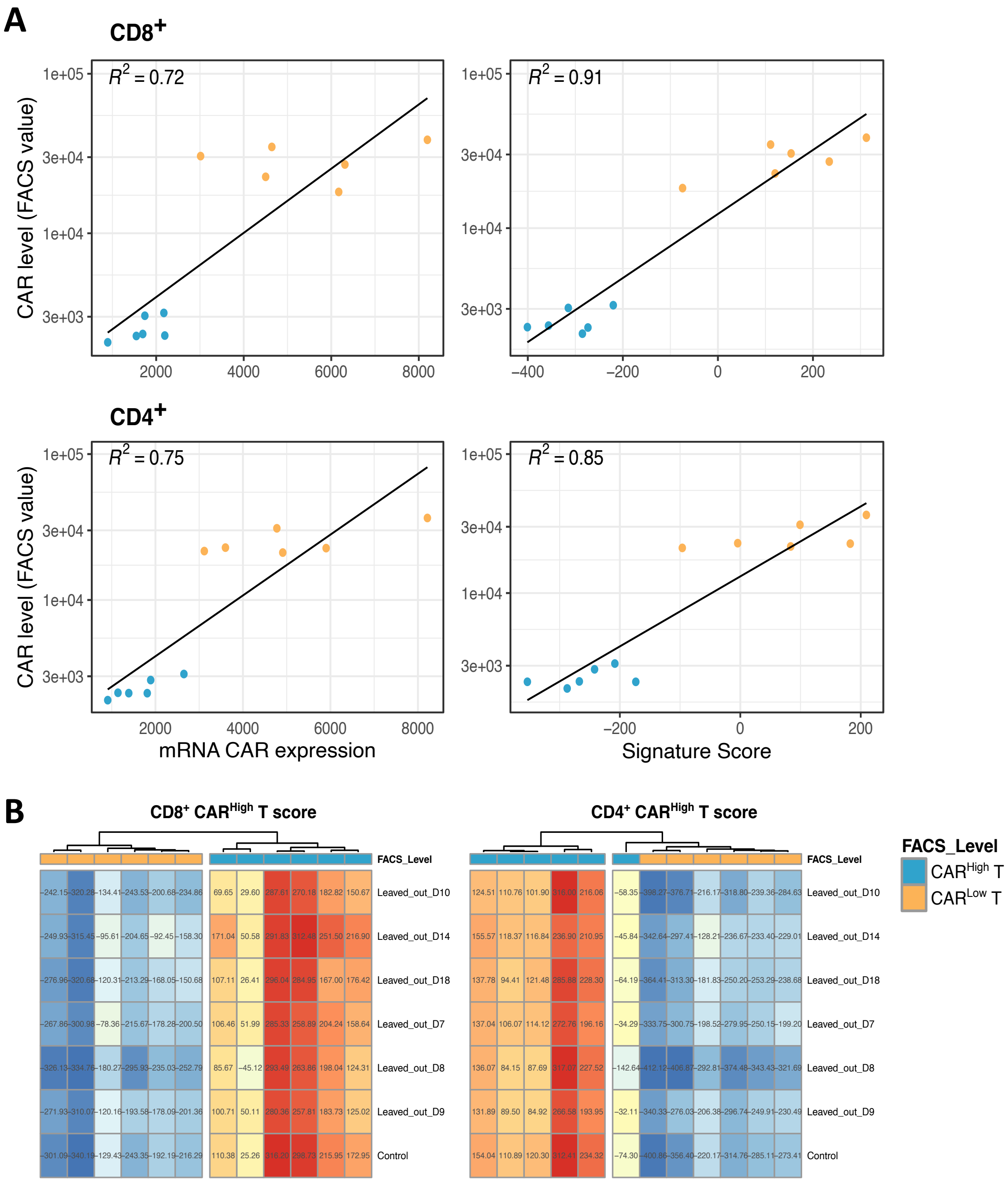


**Fig. S8. Generation and characterization of gene signatures associated to CAR$^{High}$ T cells.** Gene signatures associated to CD4$^+$ and CD8$^+$ CAR$^{High}$ T cells were developed using DEGs from bulk RNA-seq analysis between CAR$^{High}$ T and CAR$^{Low}$ T cells, as these populations have been sorted based on CAR protein expression. **(A)** Correlation between the CAR level measured by FACS and the CAR mRNA expression (left) and the developed gene signatures (right) in CD8$^+$ (upper panels) and CD4$^+$ (lower panels) cells. **(B)** Leave-one-out cross-validation of the CD8$^+$ (left) and CD4$^+$ (right) CAR$^{High}$ T signatures.

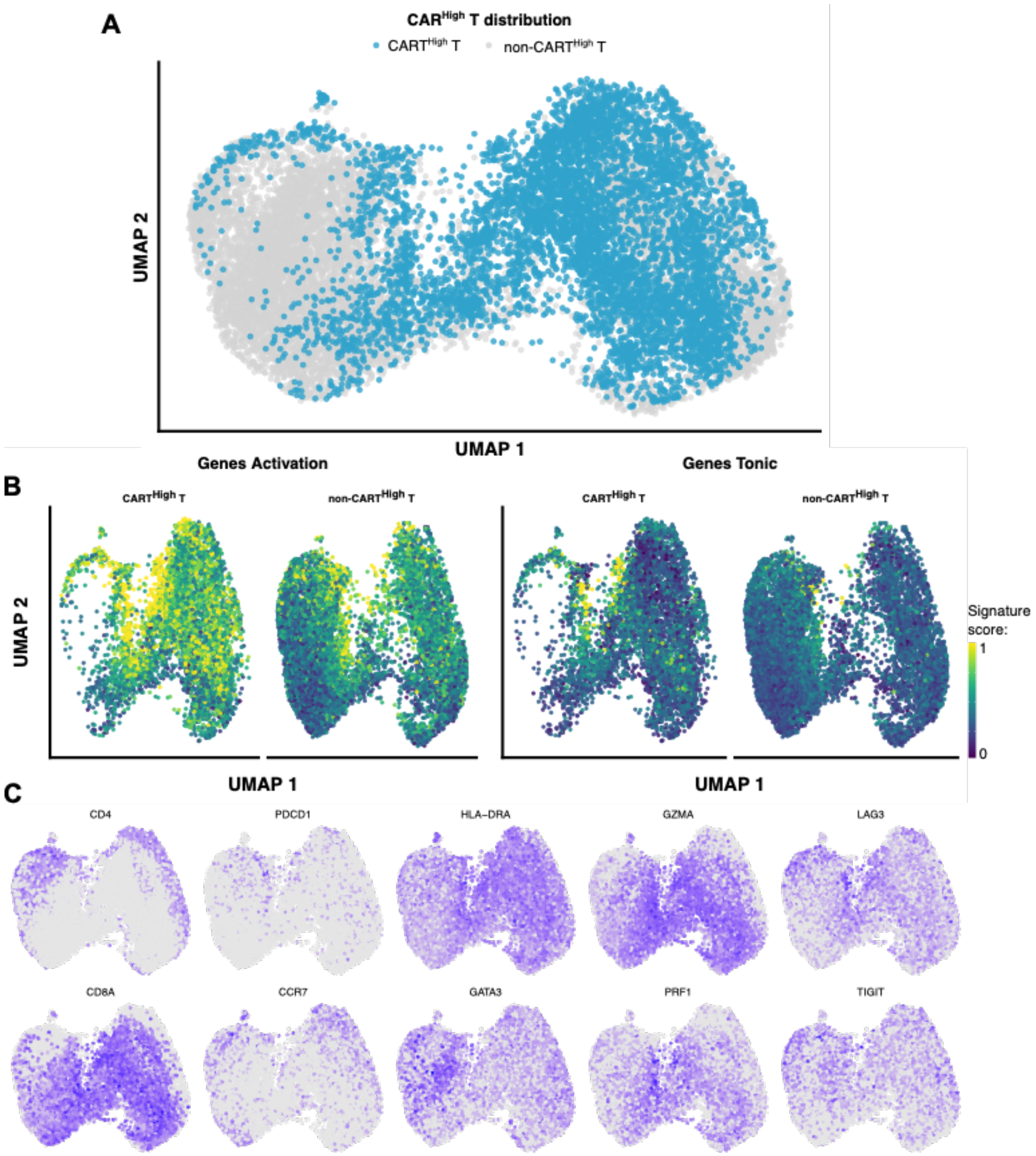


**Fig. S9. Characterization of CAR$^{High}$ T cells at single cell level.** We applied our developed signatures to an independent public dataset of CAR T products targeting CD19 to characterize CAR$^{High}$ T cells. **(A)** UMAP plot showing CAR$^{High}$ T cell distribution across analyzed cells. **(B)** UMAP plots overlaid with the score of activation and tonic signaling signatures, showing their distribution across cells. CAR$^{High}$ T cells were enriched in the score for both signatures. **(C)** UMAP plots overlaid with mRNA expression of CD4, CD8A, PDCD1, CCR7, HLA-DRA, GATA3, GZMA, PRF1, LAG3 and TIGIT.

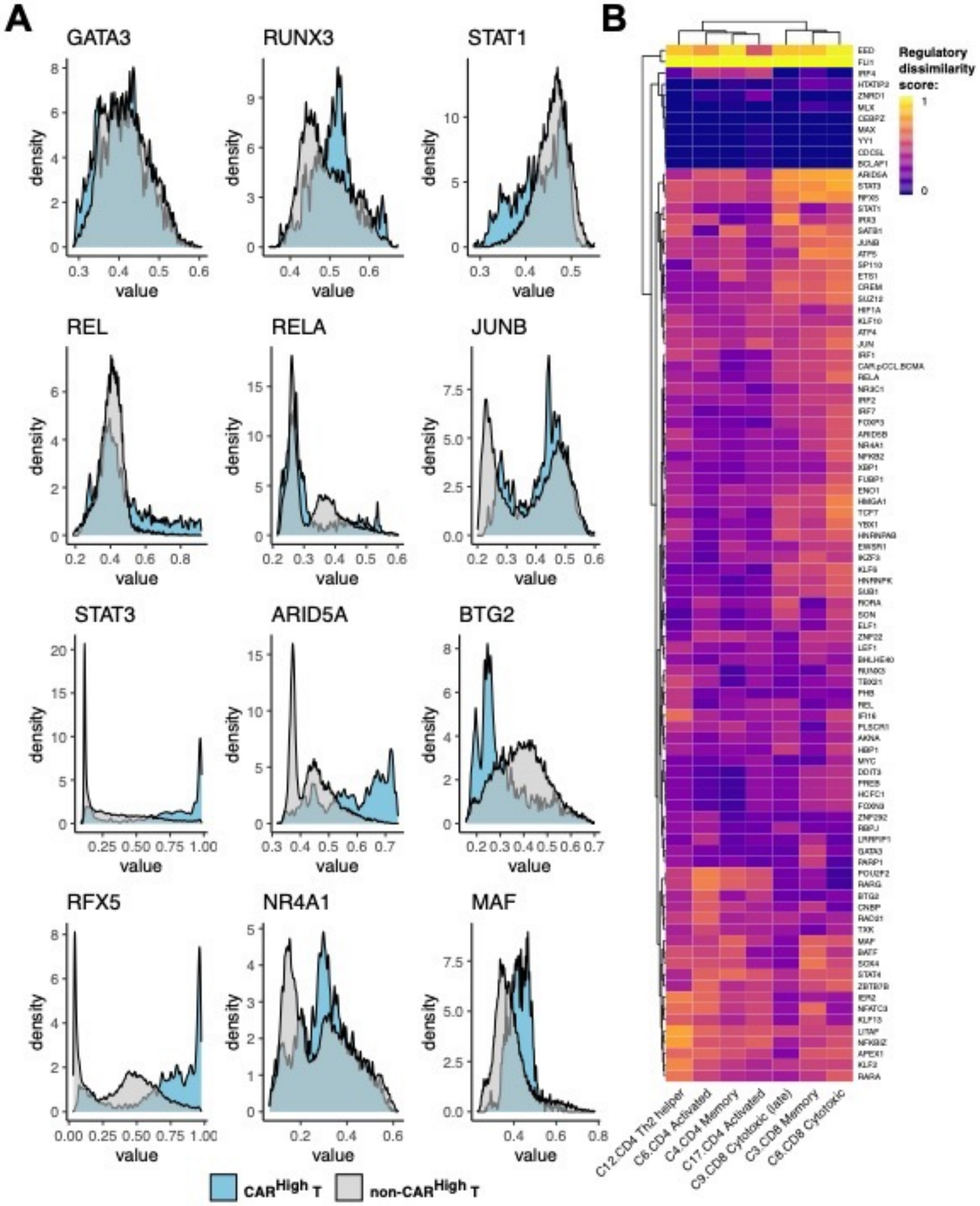


**Fig. S10. Analysis of GRNs in cell with different CAR densities.** We applied SimiC, a novel GRN inference algorithm for scRNA-seq data, to infer GRNs associated to CAR T cells with different CAR densities. **(A)** Histograms showing the activity score of GATA3, RUNX3, STAT1, REL, RELA, JUNB, STAT3 ARID5A, BTG2, RFX5, NR4A1 and MAF regulons in $CAR^{High}$ T cells versus the rest of the cells. **(B)** Heatmap showing the regulatory dissimilarity score between $CAR^{High}$ T cells and the rest of the cells of the different regulons in each cluster (provided it contain at least 5% of $CAR^{High}$ T cells).

**Legends for supplemental tables.**

**Table S1. List of differentially expressed genes in RNA-seq analysis between CAR$^{High}$ T and CAR$^{Low}$ T cells in both CD4$^{+}$ and CD8$^{+}$ cell subsets.**

Attached as separate excel file

**Table S2. List of differential peaks in ATAC-seq analysis between CAR$^{High}$ T and CAR$^{Low}$ T cells in both CD4$^{+}$ and CD8$^{+}$ cell subsets.**

Attached as separate excel file

**Table S3. List of VDJ clonotypes detected in scRNA-seq analysis of CAR T cells.**

Attached as separate excel file

**Table S4. List of the genes used for the generation of the signatures for CD4$^{+}$ and CD8$^{+}$ CAR$^{High}$ T cells.**

Attached as separate excel file

**Table S5. List of the antibodies used in this study.**

Attached as separate excel file

**Table S6. List of the primer and probe sequences used in this study.**

Attached as separate excel file

**Table S7. List of the genes used in the T cell activation and tonic signaling signatures.**

Attached as separate excel file
